# Distinct polymorphisms in HLA-class-I molecules of HbE/B-thalassemia cohort in Eastern India influences their vulnerability to Apicomplexan infections

**DOI:** 10.1101/2025.08.04.25332941

**Authors:** Shatarupa Bhattacharya, Motiur Rahaman, Shreya Suman, Deepak Kumar Rout, Shashank Purwar, Bhavna Dhingra, Praphulla Chandra Shukla, Gayatri Mukherjee, Madhulika Gupta, Mrinal Kanti Bhattacharya, Tuphan Kanti Dolai, Nishant Chakravorty, Budhaditya Mukherjee

**Affiliations:** School of Medical Science and Technology, IIT Kharagpur, India; Department of Chemistry and Chemical Biology, Indian Institute of Technology (ISM), Dhanbad, India; Department of Biochemistry, School of Life Sciences, University of Hyderabad, India; Department of Microbiology, All India Institute of Medical Sciences, Bhopal, India; Department of Pediatrics, All India Institute of Medical Sciences, Bhopal, India; Department of Hematology, Nil Ratan Sircar Medical College and Hospital, Kolkata, India

**Keywords:** Human Leukocyte Antigen, HbE/β-thalassemia, Next Generation Sequencing, Toxoplasmosis, Malaria

## Abstract

Although genetic disorders like HbE/β-thalassemia-(HBT) are associated with detrimental health outcomes, they may be subject to positive-selection in endemic regions of parasitic infections. In sub-Saharan-Africa, individuals with HBT, dependent on regular blood-transfusions have been observed to exhibit protection against malaria, while remaining susceptible to toxoplasmosis. This observation suggests, hematological genetic-disorders may significantly influence pathogenesis of Apicomplexan infections. In HBT-patients from Eastern-India, protection against toxoplasmosis was identified in individuals carrying Human-Leukocyte-Antigen-A*33-(HLA-A*33) allele. This protective effect was found to be conserved in peripheral-blood-mononuclear-cells-(PBMCs) obtained from both HBT-patients and healthy-controls. While parasite-entry into PBMCs was permitted, intracellular proliferation was restricted, which was linked to significantly enhanced CD8⁺-IFN-γ⁺ response in A*33-positive-cells relative to susceptible-genotypes. The elevated IFN-γ production was attributed to increased binding-affinity of A*33 with *Toxoplasma*-derived-peptides like SAG2C, leading to improved antigen-presentation. Interestingly, no protective role of A*33 was observed against *Plasmodium*, indicating a pathogen-specific interaction between HLA-alleles and Apicomplexan-derived-antigens. Further next-generation-sequencing of class-I and-II HLA-loci in HBT-patients revealed a potential association between HLA-C*07 and protection against *Plasmodium* infection. These findings provide mechanistic insights into how host genetic-factors may influence susceptibility to Apicomplexan infections and suggest that such interactions could contribute to evolutionary persistence of deleterious HBT-mutations in regions where these infections are co-prevalent.

**Graphical Abstract:** Graphical Abstract.
Schematic Representation of allele-pathogen dynamics explored in this study, highlighting the selective fitness advantage conferred by specific HLA variants in the context of *Toxoplasma* and *Plasmodium* infection.

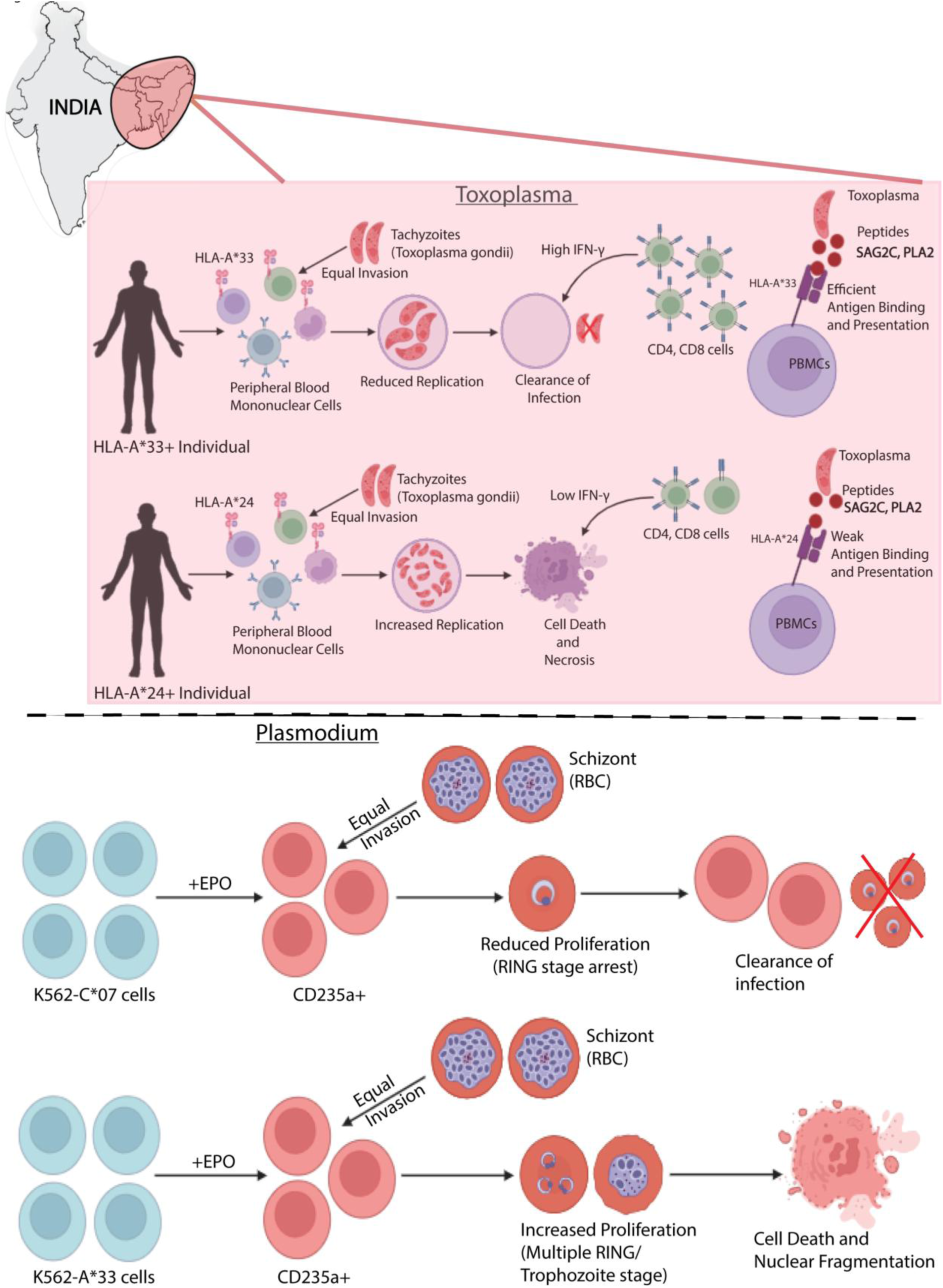

## Introduction

β-thalassemia is a prevalent monogenic disorder affecting millions worldwide, characterized by reduced or absent adult hemoglobin (HbA)[1] synthesis, resulting in chronic hemolytic anemia. An estimated 1.5% of the global population (∼80–90 million) carries the β-thalassemia trait. In India, the carrier rate is ∼2.78%[2], with substantial regional variation. India also reports high prevalence of HbE, a hemoglobin variant common in South Asia[3],[4]. Co-inheritance of HbE with β-thalassemia significantly worsens clinical outcomes, resulting in a broad phenotypic spectrum from mild to transfusion-dependent disease[5]. Many patients face lifelong transfusion needs and associated complications, including iron overload, oxidative stress, organ failure, and infection risks, particularly HIV and hepatitis, which remain major causes of mortality despite improved blood safety.

The co-endemicity of hemoglobinopathies and parasitic infections, such as *Plasmodium* and *Toxoplasma*, suggests potential host-pathogen interactions influencing disease progression[6],[7]. Hemoglobinopathies like sickle cell anemia and thalassemia have been positively selected in malaria-endemic regions due to their protective effects. Notably, HbE has been associated with reduced *Plasmodium falciparum* replication[8], implying a similar protective role. The high frequency of HbE/β-thalassemia in South-East Asia coinciding with high Apicomplexan burden highlights the need to explore host genetic factors affecting infection susceptibility. Among these, HLA alleles play a central role in modulating immune responses and have been linked to disease outcomes in infections, cancer, autoimmune disorders, and hematological conditions[9],[10],[11]. The highly polymorphic HLA region (6p21) encodes over 250 genes, including classical class I (A, B, C) and class II (DP, DQ, DR) loci. Evidence suggests HLA polymorphisms[12] influence malaria susceptibility and are shaped by local selective pressures[13],[14]. Regions where HbE mutations and infectious diseases co-occur may also drive natural selection of specific HLA alleles[15],[16].

This study investigates the interplay between HLA alleles and infection susceptibility in HbE/β-thalassemia patients from Eastern India cohort, a region burdened by both hemoglobinopathies and Apicomplexan infections. By analysing 71 HbE/β-thalassemia patients and 50 healthy controls, we identify key alleles HLA-A*33, C*07, and DQB1*03 that modulate immune responses by differentially presenting antigenic peptides of *Toxoplasma* and *Plasmodium*. These findings underscore the role of HLA-mediated CD8+ responses in shaping host resistance, offering insights into allele-specific immune protection and guiding future strategies in disease management and risk assessment.

## Methodology

### Sample Collection and Classification of the HbE/β-thalassemia patients based on disease severity

To study the phenotypic heterogeneity of the HbE/β-thalassemia, we have collected 71 HbE/β-thalassemia patient samples from N.R.S Medical College & Hospital, Kolkata under the expert supervision of physicians. Clinical and hematological data of participants were collected from medical records or by physical examination. Further, 50 healthy individual samples to serve as Control samples was also collected.

### HLA haplotying of the patients using high-resolution NGS and Sanger sequencing method

Genomic DNA was extracted from the collected blood samples using Nucleopore Genetix kit. All gDNA samples (71 HbE/β-thalassemia patients and 10 Healthy Controls) were sequenced by high resolution Illumina-sequencing method (MedGenome Labs). Five HLA loci (A, B, C, DQB1 and DRB1) were sequenced with 150X sequencing depth. Additional 40 control samples were sequenced using the bi-directional Sanger sequencing method.

### qRT-PCR and High-Resolution-Melting-Curve Analysis (HRM) to detect the presence of *Plasmodium* and *Toxoplasma* infection in HbE/β-thalassemia patient blood

Total genomic DNA extracted from blood samples as per user protocol was used as a template to detect the presence of *Plasmodium* and *Toxoplasma*. qRT-PCR-HRM assay was performed using an QuantStudio 5 real-time-PCR systems (Applied Biosystems™). The melting curve plots were generated and analysed using QuantStudio^TM^ Design & Analysis Software v1.5.2 to determine the average melting temperature (T_m_) for *Plasmodium* (Tm= 77.079 ± 1) and *Toxoplasma* (Tm= 85.247 ± 1). Fresh 10 samples were checked for parasite presence using the same method, and the HLA allelic information was confirmed using PCR. The primer sequences for RT-qPCR are enlisted in **Table 1**.

**Table 1.**
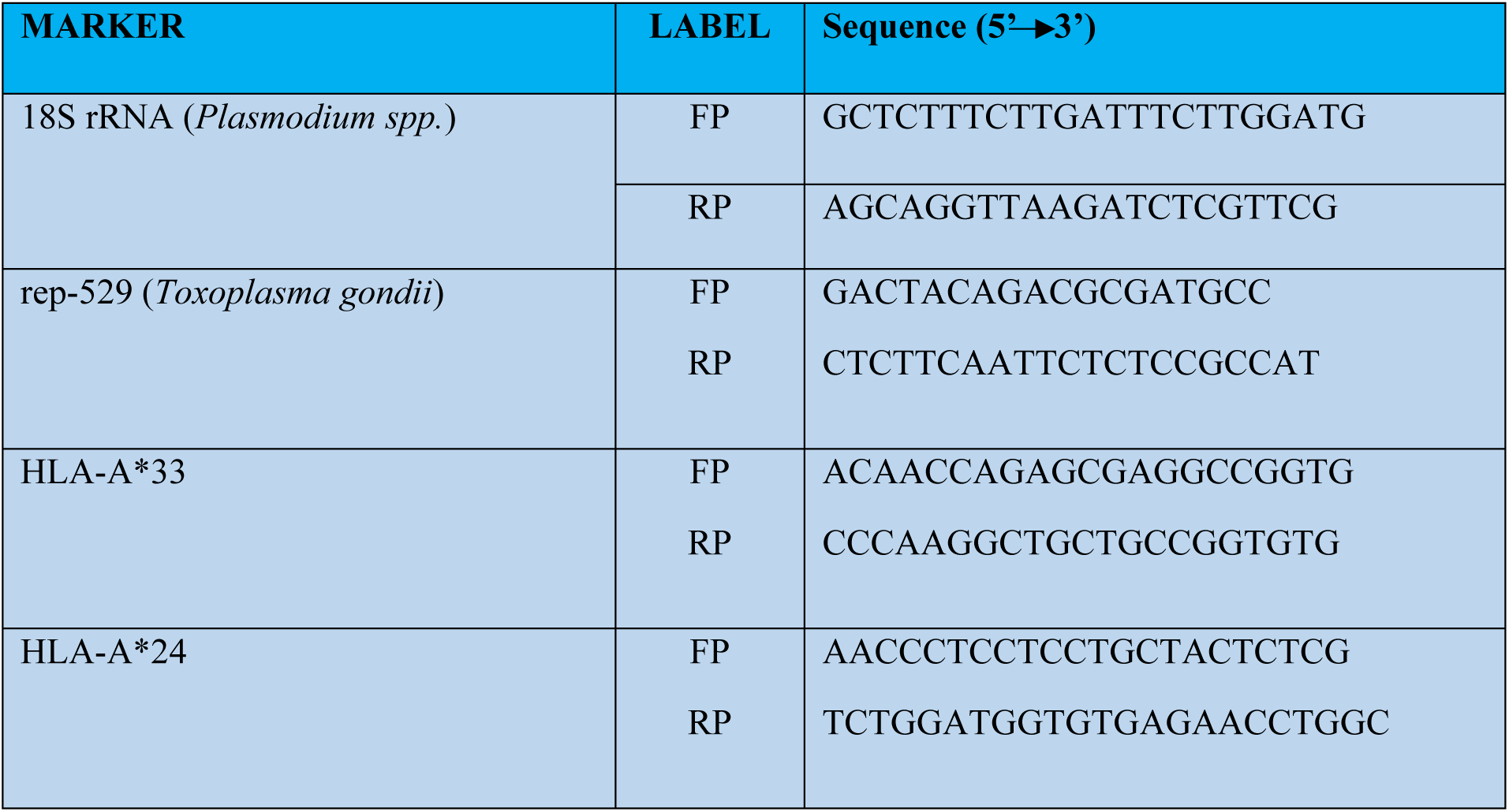
Oligos for Apicomplexan infection and HLA detection via q RT-PCR.

### Cell Culture

Confluent monolayers of human foreskin fibroblasts (HFFs), were cultured in Roswell Park Memorial Institute (RPMI) 1640 medium (GIBCO), supplemented with 10% FBS (GIBCO) and 100 U PenStrep/Gentamycin (GIBCO). The cells were left to adhere for 24h at 37^0^C and 5% CO_2_ before infection. K562 cells (HLA-null)[17],[18] were used for the HLA overexpressing studies as previously reported[19],[20]. They were routinely cultured in the similar method, as mentioned above. Isolated PBMCs[21] were also cultured in the similar method.

### *Toxoplasma* Culture and Ex-vivo Infection model

*Toxoplasma gondii* (ΔKu80) and GFP-ΔKu80 were maintained in the same conditions as mentioned above. PBMCs from the HbE/β-thalassemia patients/healthy controls were infected with freshly egressed tachyzoites at a MOI of 1:10, and used for respective experiments.

### *Plasmodium* culture and Synchronization

*Plasmodium falciparum* 3D7 cultures were maintained by the candle jar method at 37°C as described previously at 5% haematocrit, with 0.5% albumax (Invitrogen, Carlsbad, CA, USA) and 0.005% hypoxanthine supplemented RPMI 1640 medium. Parasite cultures were synchronized at the ring stage by treatment with 5% sorbitol (Sigma, St. Louis, MO, USA).

### In vitro Infection

For in vitro infection, K562-HLA-A*33/24 cells were infected with freshly egressed tachyzoites at a MOI of 1:10 for different time points, as per the specific experimental requirement.

For in vitro infection, K562-HLA-C*07 cells were infected with RBCs containing *P. falciparum* 3D7 parasites (late schizonts) at a 5% parasitemia, for 6h, 12h, 24h, 36h and 48h, thereafter fixed in Methanol and stained with Geimsa stain accordingly, to count the total number of infected cells and calculate the infectivity index.

### Red/Green Invasion Assay

Freshly egressed tachyzoites were used to infect the A*33, A*24 PBMCs to assess the early invasion by the parasites. The infected cells were spun down at 1000rpm for 2 minutes, to facilitate closer interaction of the parasites with cells. The cells were then kept in the 37℃-water bath for 15 minutes. Next the cells were transferred to the CO_2_ incubator and kept there for 35 mins. Subsequently the cells were fixed in 4% Paraformaldehyde /0.05% Glutaraldehyde for 5 minutes, and taken for microscopy.

### Immunofluorescence Assay (IFA) and Confocal Microscopy

For IFA, the following antibodies were used - rabbit anti-GAP45 and anti-SAG1. All images were captured using Olympus confocal microscopy (FV3000) and Leica Stellaris 5 Dmi8 demonstration unit. Final image analysis and processing were done with ImageJ.

### Cytokine Measurement

IFN-ℽ levels were measured in the infected A*33, A*24 PBMCs supernatant, at different time points post-infection, by ELISA (R&D Systems) following the manufacturer’s manual.

### Immunostaining and Flow-Cytometry based cell binding assay

All flow-cytometric experiments were carried out in a BD-LSRFortessa™ instrument, and analyses were done using FlowJo™ software version 10.6.0 (Becton, Dickinson and Company, Ashland, OR; 2019). To study proliferation of MHC-I expressing cells, and CD8+and CD4+ T-cells, isolated PBMCs (A*33+ and A*33-) infected with Tachyzoites, were stained with anti-CD4 (PerCP-Cy5.5/APC) (BioLegend) (1:100 dilution), anti-CD8 (FITC) (Thermo) (1:50 dilution) and anti-MHC-I (FITC) (Abclonal) (1:100 dilution) antibodies. Cells were then washed, fixed, and analyzed by flow-cytometry for binding of MHC-I, CD4+ and CD8+ T-cells.

### Apoptosis Assay

Infection mediated apoptosis in PBMCs (A*33+, A*33-) was detected by Annexin V Conjugates for Apoptosis Detection staining (Invitrogen). The infected cells were also Giemsa stained and counted at 10X resolution using the Leica bright field microscope to assess the number of viable cells left.

### Induction of Differentiation in K562 cells to serve as model for *P. falciparum* infection

Erythroid differentiation was performed in the presence of Recombinant-human-EPO (3IU/ml) for 72h. Post differentiation, FACS was carried out to check the expression of CD235a (Glycophorin-A), which is essential for the entry of *P. falciparum* parasites into the cells.

### Overexpression of HLA-C*07 allele in K562 cells

pcDNA3.1/C*07 construct was transfected into K562 cells using Lipofectamine™ 2000(Invitrogen) and screened with 1000µg/ml G418 disulfide(A1720-Sigma) for over 7 days. The positive clones were confirmed by RT-qPCR and flow-cytometry with anti-MHC-I antibody for C*07 surface expression.

### Cloning and over-expression of mCherry-A*33 and mCherry-A*24 alleles

The full length of mCherry was amplified from pcDNA3.1- and cloned into the A*33- pcDNA3.1 backbone plasmids using the NEB-Gibson Assembly Master-Mix(Lot #10229799). Positive clones were confirmed by RT-qPCR, IFA and immunoblot for mcherry-HLA-A*33/24 expression.

### Cloning and purification of C-terminal His-tg-A*33 and His-tg-A*24 proteins

The full length of 6X His-Tg was amplified from the 2.8-lic-HisTag plasmid and cloned into the A*33, A*24 backbone plasmid using the NEB-Gibson Assembly Master Mix. His-Tg/HLA construct was purified from the K562 cells, as per previously used protocols[22],[23]. The purified protein fractions were then run on a 12% SDS gel to assess their purity, and further protein expression was confirmed by immunoblotting using anti-HisTg antibody.

### Immunoblotting analysis

Immunoblot directed against (anti-MHC-Class-I-(EMR8-5) mouse mAb #88274, anti-mCherry rabbit polyclonal antibody, anti-His-Tag rabbit (D3I1O) XP) (Cell-signalling technology) was performed as per previously reported protocol[24].

### Structural Modelling of A*33 and Peptide Selection for Docking Studies

Since MHC-Class-I-(HLA I) Heavy Chain α covers the entire binding groove domain[25],[26] and IMGT/HLA database primarily contains Chain A’s exact sequences, it is the only chain considered for the alignment and structure prediction using Alphafold2, as previously reported[27],[28],[29]. A total of 15 peptides derived from the protozoan parasite *Toxoplasma* were docked with both the A*33 and the A*24 using HADDOCK. The binding interactions of 2 peptides (PLA2, SAG2C) with A*33 and A*24 were analysed using Discovery-Studio 2024 (BIOVIA DS, San Diego:Dassault Systèmes, 2024) to identify key residues involved in binding to assess stability of the complexes. These key amino acids were mutated by the other 19 amino acids and modelled using Alphafold2. These mutated peptides were again docked with the HLA-A*33 and A*24 using HADDOCK with the same protocol discussed above.

### Surface plasmon resonance for protein-peptide interaction study

The interaction of A*33, A*24 with wild-type/mutant SAG2C was analyzed using Biacore-T200. A*33 protein was immobilized on a CM5 chip (Cytiva) and different concentrations of SAG2C as an analyte (1–200 μM) were injected at a flow rate of 30μl/min for 120s. The results were analyzed using BIA-evaluation-Software 2.0 and Graph Pad Prism 8.0.

### Statistical Analysis

The statistical analysis was carried out using GraphPad Prism 8 software. The results were presented as Mean ±SD and one-way ANOVA was performed to compare the study groups. Odds ratio (ORs) and 95% confidence intervals were calculated as statistical differences regarding HLA frequencies. p-values <0.05 were considered to be statistically significant. All experiments were repeated at least three times and in triplicates. The comparison between two groups were performed by two-tailed Student’s t-test.

## Results

### *Toxoplasma and Plasmodium* Infection positivity among HbE/β-thalassemia patients is independent of blood transfusion

To investigate the potential occurrence of Apicomplexan infection in relation to transfusion dependency, 71 HbE/β-thalassemia (HBT) patients were recruited. Based on disease severity and transfusion requirements[30], 15 patients were classified as transfusion-dependent thalassemia (TDT), and 46 as non-transfusion-dependent thalassemia (NTDT). High-resolution melting (HRM) curve analysis was used to detect *Plasmodium* and *Toxoplasma* DNA in patient blood (**Figure 1.A.i-ii.**). Among 71 asymptomatic HBT patients, 29 (40.84%) were *Plasmodium*-positive and 14 (19.71%) were *Toxoplasma*-positive, whereas none of the 50 healthy controls (HC) were positive (**Figure 1.B.i-ii.**). Co-infection with both pathogens was rare (n=3), confirming assay specificity. Surprisingly, infection prevalence did not correlate with transfusion dependence (**Figure 1.C.i-ii.**). Instead, *Plasmodium* positivity was significantly higher in NTDT patients (**Figure 1.C.ii.)**, suggesting additional host factors may influence susceptibility. Given previous associations between HLA polymorphisms, transfusion-related outcomes, and parasitic susceptibility, next-generation sequencing (NGS) of class I/II HLA loci was performed on 61 QC-passed HBT samples and 10 HC samples, identifying 92 alleles (**Figure 1.D.**) (**Supplementary Table 1**). Five alleles (A*33, C*07, DQB1*03, DQB1*02, DRB1*07) were shortlisted based on occurrence in ≥33% of the cohort (p < 0.0001), which were further validated in 40 additional HC samples via Sanger sequencing (**Figure 1.E.**) (**Supplementary Table 2**). Among the top hits, HLA-A*33 showed a significant negative association with *Toxoplasma* infection. Among A*33-positive individuals, only 27.28% were *Toxoplasma*-positive, while 51.51% were *Toxoplasma*-negative HBT patients, and 21.21% were HC. A*33 carriers were significantly more likely to be *Toxoplasma*-negative [OR = 0.51, CI = 0.30–0.90, p = 0.02] and showed enrichment compared to HC [OR = 2.47, CI = 1.39–4.35, p = 0.002] (**Figure 1.F.i.**), indicating a protective role of A*33.

**Figure 1.**
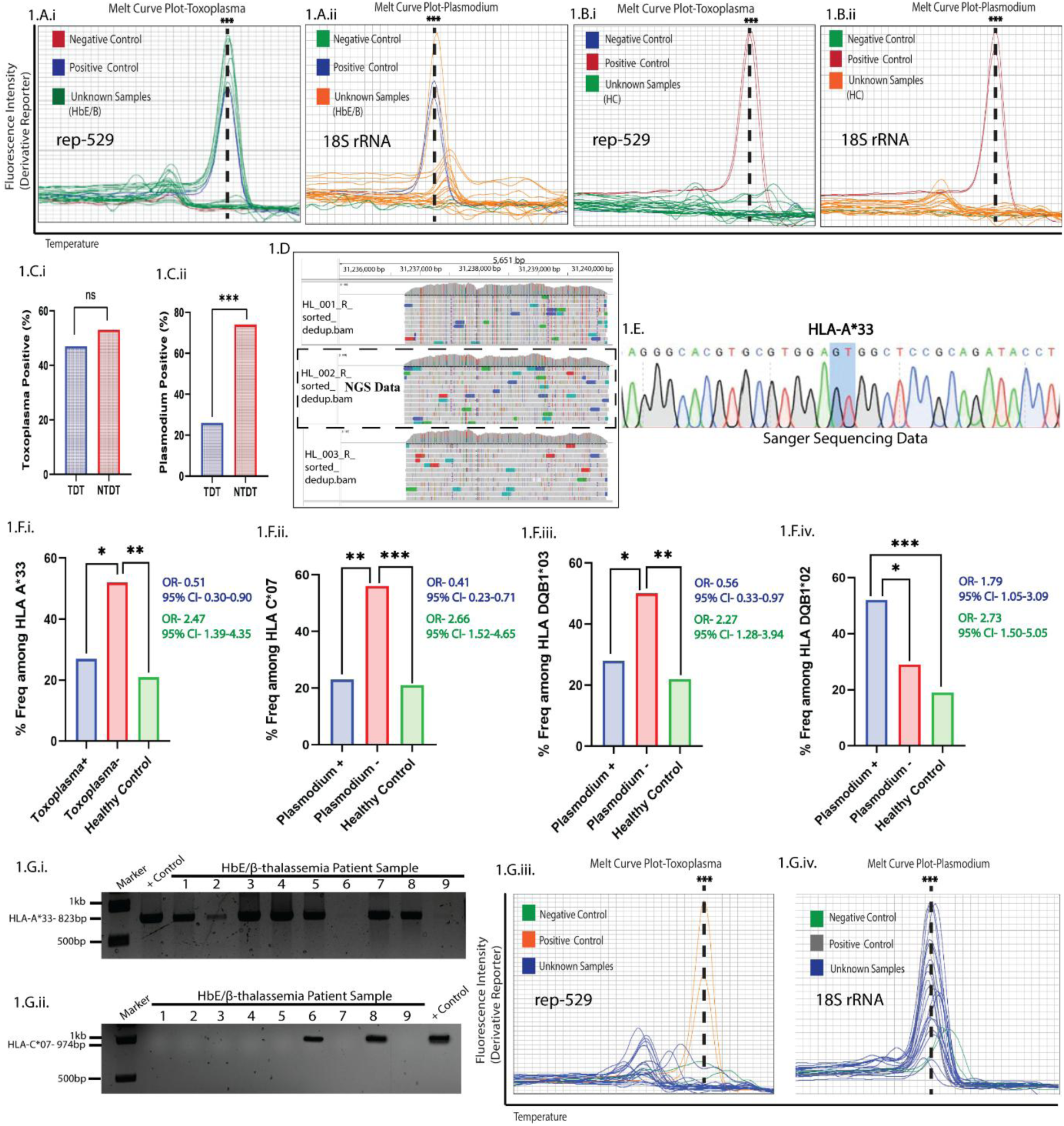
Transfusion dependence is not linked with higher prevalence of *Toxoplasma/Plasmodium* infection among HbE/β-thalassemia patient. **(A.i.-B.i.)** qRT-PCR based HRM curve analysis for *Toxoplasma* rep-529 detection in HbE/β-thalassemia patients (n=61) and Healthy Controls (n=50), respectively. **(A.ii.-B.ii)** qRT-PCR based HRM curve analysis for *Plasmodium* 18S rRNA detection in HbE/β-thalassemia patients (n=61) and Healthy Controls (n=50), respectively. **(C.i-ii.)** Co-relation analysis between *Toxoplasma* and *Plasmodium* infection positivity with Transfusion Dependency in HbE/β-thalassemia patients. **(D.)** NGS (150X) based raw sequence read of HbE/β-thalassemia patients (HL_001, HL_002 and HL_003) aligned using a FASTA-like algorithm. The dotted box indicates presence of HLA-A*33 in HbE/β-thalassemia patient sample HL_002. **(E.)** Representative Sanger Sequence Chromatogram of Healthy Control DNA denoting HLA-A*33. The red box marks the position of the substitution. **(F.i-iii.)** Percentage frequency of HLA alleles (A*33, C*07, DQB1*03) from *Toxoplasma* and *Plasmodium* infection, respectively. **(F.iv.)** Percentage frequency of HLA allele (DQB1*02) showing susceptibility towards *Plasmodium* infection. **(G.i-ii.)** Genomic DNA PCR based detection of protective HLA alleles (A*33, C*07) in HbE/β-thalassemia patients (n=9). (**G.iii.)** qRT-PCR based HRM curve analysis for *Toxoplasma* rep-529 detection in HbE/β-thalassemia patients (n=9). (**G.iv.**) qRT-PCR based HRM curve analysis for *Plasmodium* 18S rRNA detection in HbE/β-thalassemia patients (n=9). p ≤ 0.05 is marked as *, p ≤ 0.01 is marked as ** and p ≤ 0.001 is marked as ***.

For *Plasmodium*, protective associations were identified with HLA-C*07 and HLA-DQB1*03. Among C*07 carriers, 56.41% were *Plasmodium*-negative, 23.08% were positive, and 20.51% were HC. *Plasmodium* infection was less likely among C*07 carriers [OR = 0.41, CI = 0.23–0.71, p = 0.002] versus HC [OR = 2.66, CI = 1.52–4.65, p = 0.0007] (**Figure 1.F.ii.**). Similarly, DQB1*03 carriers were significantly more likely to be *Plasmodium*-negative [OR = 0.56, CI = 0.33–0.97, p = 0.04] and enriched relative to HC [OR = 2.27, CI = 1.28–3.94, p = 0.005] (**Figure 1.F.iii.**). In contrast, HLA-DQB1*02 was positively associated with *Plasmodium* infection [OR = 1.79, CI = 1.05–3.09, p = 0.03] (**Figure 1.F.iv.**). To validate these associations, 10 new HBT samples were tested. Seven A*33-positive samples (**Figure 1.G.i.**) were all *Toxoplasma*-negative (**Figure 1.G.iii.**), while both C*07-positive (**Figure 1.G.ii.**) samples were *Plasmodium*-negative. Of the seven C*07-negative samples, five were *Plasmodium*-positive (**Figure 1.G.iv.**), reinforcing the potential protective roles of A*33 and C*07. These findings suggest specific HLA alleles may modulate susceptibility to Apicomplexan infections in HBT patients, independent of transfusion dependency, providing insight into host genetic contributions to infection outcomes.

### A*33 restricts intracelluar proliferation of *Toxoplama* leading to survival of infected host

To evaluate the role of HLA-A*33 in restricting *Toxoplasma* infection, ex vivo assays were performed using PBMCs from HbE/β-thalassemia patients with A*33+ and A*24+ (most prevalent allele in our *Toxoplasma* infected cohort) genotypes. A*24+ PBMCs showed a progressive increase in intracellular tachyzoite load, while A*33+ cells exhibited a sharp decline in parasite count post 24h, indicating effective restriction of replication and clearance (**Figure 2.A.i–iii.**). Similar assays in healthy A*33+ and A*24+ PBMCs confirmed this protective phenotype (**Figure 2.B.i–iii.**). To determine whether reduced tachyzoite burden in A*33+ PBMCs was due to impaired invasion (**Figure 2.C.i-ii.**), invasion assays were conducted (**Figure 2.C.iii.)**. Comparable levels of intracellular (red-GAP45) and extracellular (green-SAG1) parasites were observed in A*33+ and A*24+ PBMCs from both patients and healthy individuals, indicating no significant difference in invasion efficiency (**Figure 2.D.i– iv.**). Further, significantly declined tachyzoites per parasitophorous vacuole in A*33+ PBMCs were observed between 24h and 72h P.I., A*24+ PBMCs showed increased parasite proliferation with large PV showing 16 or more tachyzoites (**Figure 2.E.i–ii.**). Finally, HLA-null K562-cells expressing mCherry-tagged A*33 or A*24 (**Supplementary** Figure 1**.A., B.i-ii., C.)** confirmed these findings. A*33-K562 displayed significantly reduced parasite load at all-time points (**Figure 2.F.i–iii., 2.G.i–iii.**), invasion efficiency remained comparable for A*33-K562 and A*24-K562 (**Supplementary** Figure 1**.D.i-ii.**). Collectively, these results demonstrate that A*33 confers a conserved protective effect against *Toxoplasma* by restricting intracellular replication, not invasion. Apart from limiting tachyzoite proliferation, A*33+ PBMCs remained intact till 120h P.I., unlike A*24+ PBMCs, which showed gradual lysis 72h P.I (**Supplementary** Figure 1**.E.i-ii.**). Annexin V-FITC based apoptosis assay revealed higher late apoptosis (68.6%) and necrosis (11.6%) in A*24+ PBMCs vs. A*33+ at 48h–72h P.I. (**Figure 3.A–D.**), indicating A*33 allele protects against *Toxoplasma*-induced apoptosis[31],[32],[33].

**Figure 2.**
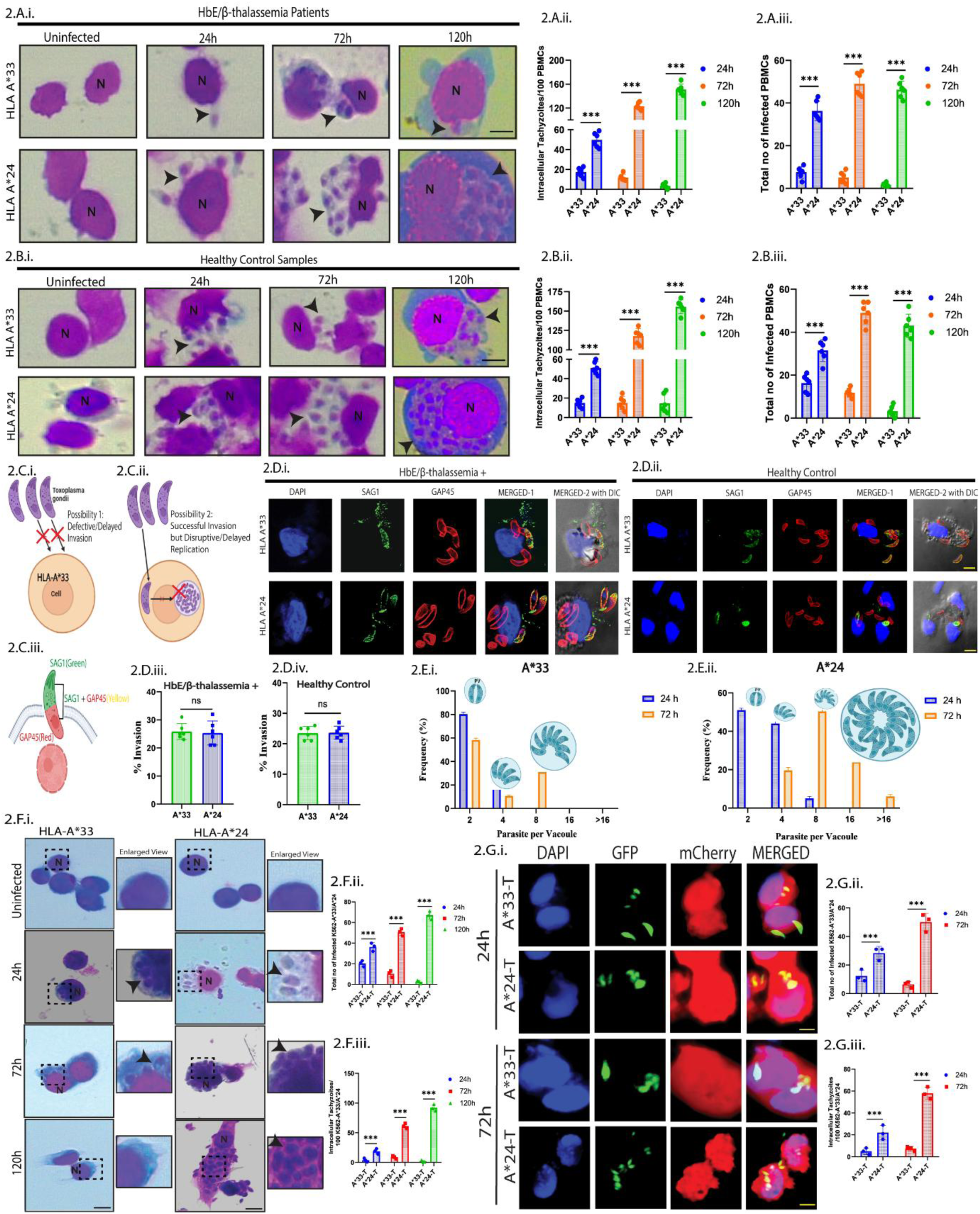
A*33 hinders successful intracellular replication of *Toxoplasma* tachyzoites. **(A.i.)** Representative Giemsa stain images of A*33 and A*24 infected HbE/β-thalassemia patient PBMCs at 24h, 72h and 120h P.I. Black arrow represents *Toxoplasma* tachyzoites. N denotes host nucleus. Scale bar 10μM. (**A.ii-iii.**) Intracellular *Toxoplasma* tachyzoite count and Infectivity Index for 24h, 72h and 120h P.I. Each dot represents count from 100 infected HbE/β-thalassemia patient PBMCs (n=6). (**B.i**) Representative Giemsa stain images of infected A*33 and A*24 Healthy Control PBMCs at 24h, 72h and 120h P.I. Black arrow represents *Toxoplasma* tachyzoites. N denotes host nucleus. Scale bar 10μM. (**B.ii-iii.**) Intracellular *Toxoplasma* tachyzoite count and Infectivity Index for 24h, 72h and 120h P.I. Each dot represents count from 100 infected Healthy Control PBMCs (n=6). (**C.i-ii.**) Scheme showing different possibilities of *Toxoplasma* tachyzoite infection in PBMCs (i) Defective/Delayed Invasion (ii) Successful Invasion but Disruptive/Delayed Replication. (**C.iii.**) Schematic representation of differential antibody staining based Invasion assay: SAG1-Extracellular (Green) GAP45-Intracellular (Red) SAG1+GAP45-Extracellular (Yellow). (**D.i.**) Representative confocal images of HbE/β-thalassemia patient PBMCs infected with *Toxoplasma* tachyzoites using a red/green invasion assay. External tachyzoites are stained green, internal tachyzoites are stained red, host nuclei are DAPI stained-blue. (**D.ii.**) Representative confocal images of Healthy control PBMCs infected with *Toxoplasma* tachyzoites using a red/green invasion assay. External tachyzoites are stained green, internal tachyzoites are stained red and host nuclei are DAPI stained-blue. (**D.iii-iv.**) Quantification of % invasion (Total Red tachyzoites/Total Green tachyzoites*100) using the red/green assay. Data is represented as Mean ± SD from 3 independent experiments. (**E.i-ii.**) Growth kinetics of *Toxoplasma* tachyzoites in A*33 and A*24 PBMCs evaluated by counting number of parasites per parasitophorous vacuole (PVs). Percentage of PVs with 2, 4, 8, 16 and >16 parasites at 24h, 72h P.I. was plotted. (**F.i.**) Representative Giemsa stain images of K562- A*33 (T) and K562-A*24 (T) infected cells at 24h, 72h and 120h P.I. Black arrow represents *Toxoplasma* tachyzoites. N denotes host nucleus. Scale bar 10μM. Enlarged view of F.i. showing infected cells. (**F.ii-iii.**) Infectivity Index and Intracellular *Toxoplasma* tachyzoite count for 24h, 72h and 120h P.I. Each dot represents count from 100 infected A*33 (T) and A*24 (T) cells (n=3). (**G.i.**) Confocal images of GFP-tagged *Toxoplasma* tachyzoite infected mCherry-K562-A*33 (T) and mCherry-K562-A*24 (T) cells at 24h and 72h P.I. Scale bar 10μM. (**G.ii-iii.**) Infectivity Index and Intracellular *Toxoplasma* tachyzoite count for 24h and 72h P.I. Each dot represents count from 100 infected A*33 (T) and A*24 (T) cells (n=3). Each experiment was performed in 3 biological replicates and graphical data are represented as Mean ± SD. p ≤ 0.05 is marked as *, p ≤ 0.01 is marked as ** and p ≤ 0.001 is marked as ***.

**Figure 3.**
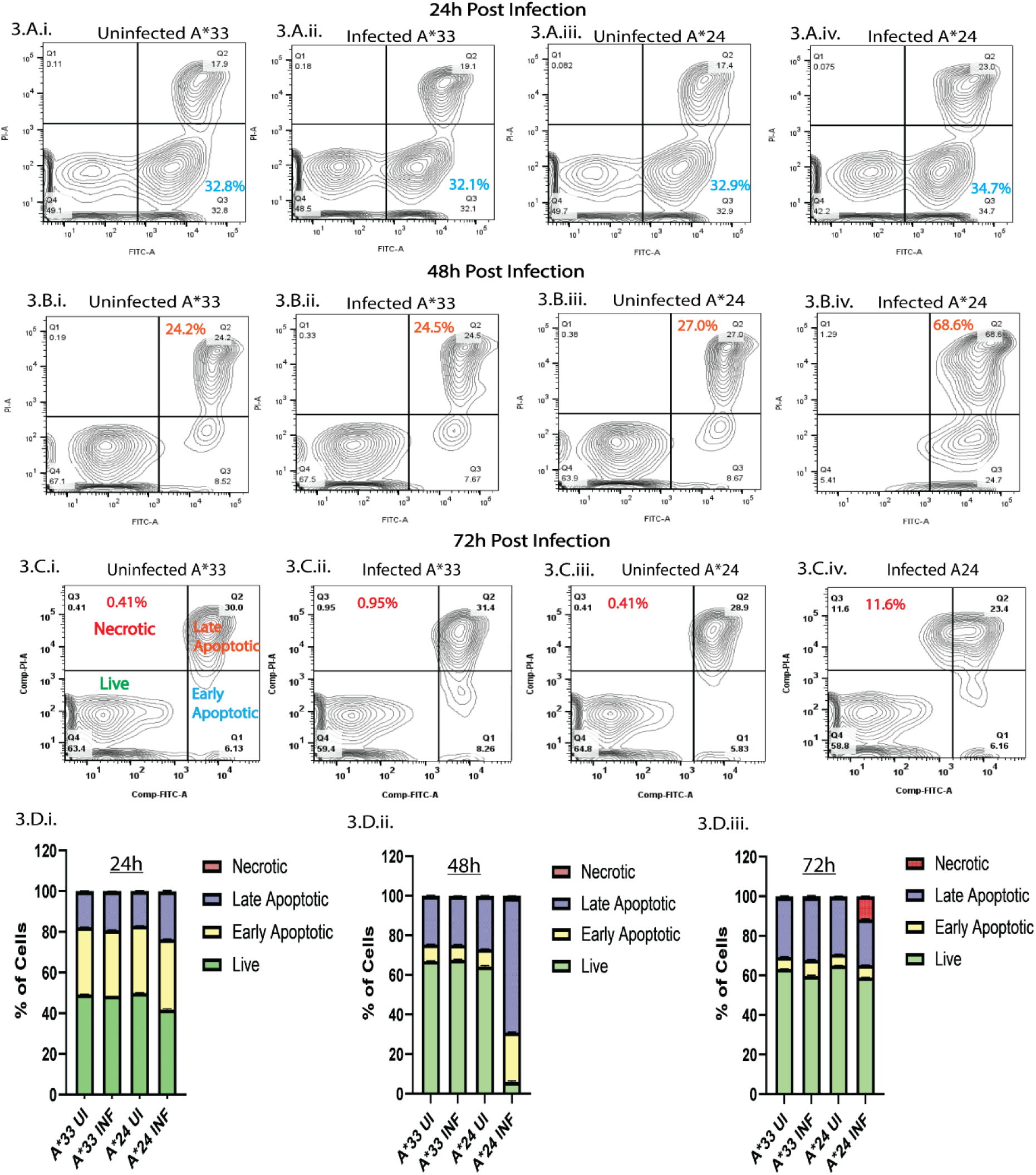
*Toxoplasma* infected A*24 PBMCs undergoes Apoptosis. (**A.i-iv.**) Contour Plot representing Flow cytometry evaluation depicting early apoptosis in different groups; A*33 UI, A*33 I; A*24 UI, A*24 I (from left to right) 24h P.I. (**B.i-iv.**) Contour Plot representing Flow cytometry evaluation depicting late apoptosis in different groups; A*33 UI, A*33 I; A*24 UI, A*24 I (from left to right) 48h P.I. (**C.i-iv.**) Contour Plot representing Flow cytometry evaluation depicting necrosis in different groups; A*33 UI, A*33 I; A*24 UI, A*24 I (from left to right) 72h P.I. (**D.i-iii.**) The percentage (%) of live, early apoptotic, late apoptotic and necrotic PBMCs after *Toxoplasma* infection between the aforementioned groups; 24h, 48h and 72h P.I. respectively (from left to right). Each experiment was performed in 3 biological replicates and graphical data are represented as Mean ± SEM.

### A*33 is linked with increased MHC-I presentation and CD8 mediated IFN-ℽ response in *Toxoplasma* infected host

Since A*33 restricted tachyzoite replication and apoptosis, and CD8-dependent IFN-γ is known to limit intracellular *Toxoplasma* proliferation[34],[35],[36],[37],[38], it was hypothesized that A*33 enhances antigen presentation, triggering a stronger CD8-mediated IFN-γ response. IFN-γ levels were measured in supernatants of infected A*33 and A*24 PBMCs (**Figure 4.A.**). A*33 PBMCs showed a steady increase in IFN-γ, peaking before it plateaued around 120h, likely due to parasite clearance. In contrast, A*24 PBMCs showed persistently low IFN-γ (**Figure 4.A.**). This response aligned with MHC-I expression: while almost similar at 24h (**Figure 4.B.i.**), A*33 PBMCs showed significantly increased MHC-I at 72h (**Figure 4.B.ii.**), unlike A*24 PBMCs (**Figure 4.B.iii.**). Additionally, A*33 PBMCs exhibited higher CD8 and CD4 activation at 72h (**Figure 4.C.i-ii., D.i-ii.**). Notably, CD8+ T-cells, key IFN-γ producers, were more abundant than CD4+ subset in A*33+ PBMCs at 72h (**Figure 4.E.**), supporting the conclusion that A*33 promotes CD8-driven IFN-γ responses through enhanced MHC-I antigen presentation, aiding in effective parasite clearance.

**Figure 4.**
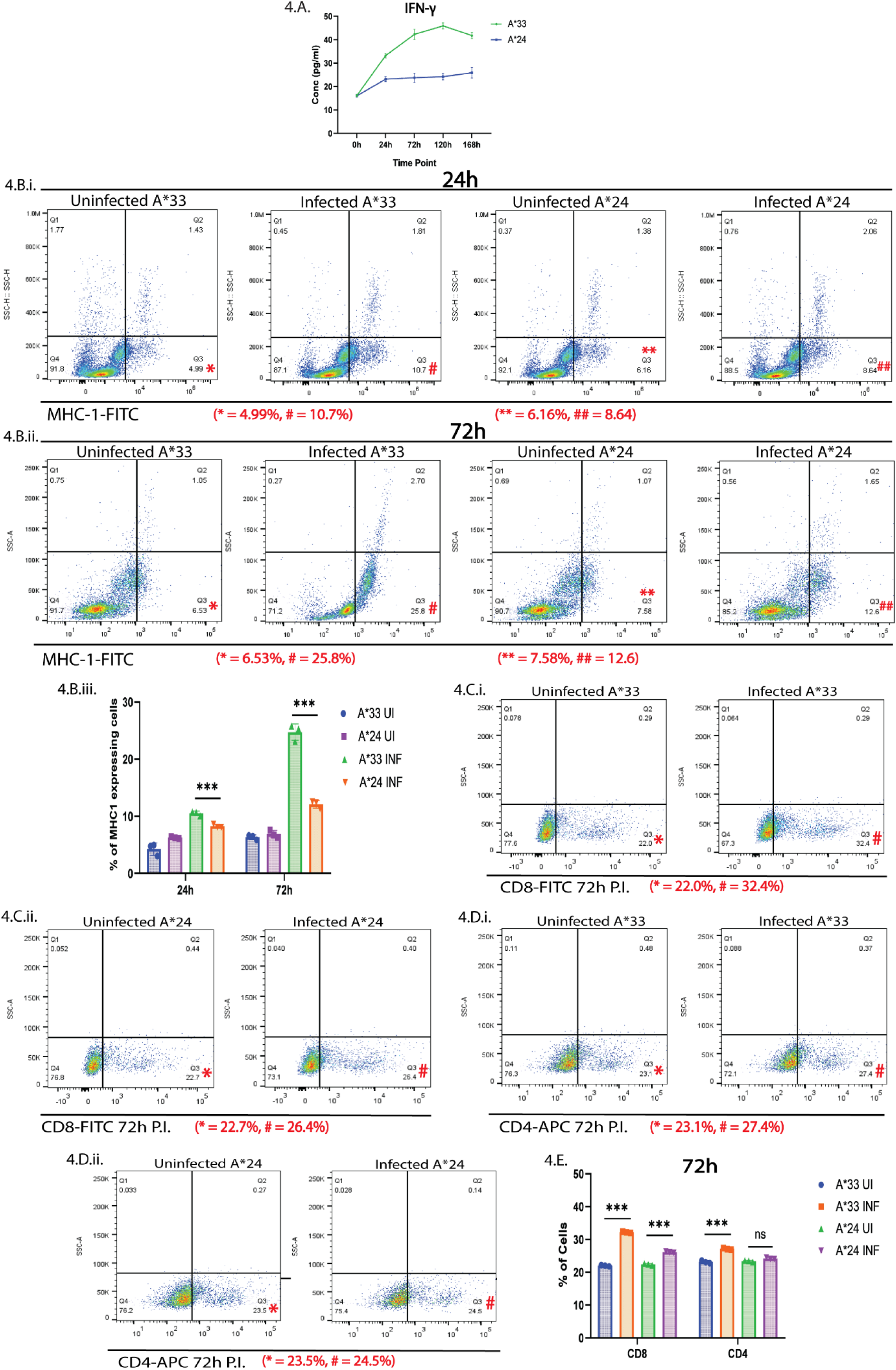
*Toxoplasma* infected A*33 PBMCs show significantly high MHC-I expression and CD8 dependent IFN-γ mediated pro-inflammatory response. **(A.)** IFN-γ level in the supernatant of *Toxoplasma* infected A*33 and A*24 PBMCs 24h, 72h, 120h and 168h P.I. Data is represented as Mean ± SD from 3 independent experiments. (**B.i-ii.**) Representative Flow cytometry-based scatter plot showing enrichment of FITC-MHC-I expressing cells in Infected A*33 and A*24 PBMCs in comparison to the uninfected control group, 24h and 72h P.I., respectively (**B.iii.**) Bar graph representing Flow cytometry data from 24h and 72h P.I., respectively, as in B.i., B.ii. are represented. (**C.i-ii.**) Representative Flow cytometry-based scatter plot showing enrichment of FITC-CD8+ T cells in Infected A*33 and A*24 PBMCs in comparison to the uninfected control group, 72h P.I. (**D.i-ii.**) Representative Flow cytometry-based scatter plot showing enrichment of APC-CD4+ T cells in Infected A*33 and A*24 PBMCs in comparison to the uninfected control group, 72h P.I. (**E.**) Bar graph representing Flow cytometry data from 72h P.I., as in C(i-ii), D(i-ii) are represented. Data is represented as Mean ± SD from 3 independent experiments. p ≤ 0.05 is marked as *, p ≤ 0.01 is marked as ** and p ≤ 0.001 is marked as ***.

### Ser13 residue in *Toxoplasma* Antigenic peptide SAG2C interacts with antigen binding pocket of A*33

Since A*33 (protective) and A*24 (susceptible) show differential MHC-I expression during *Toxoplasma* infection and MHC-I stability depends on peptide binding, it is likely that A*33 presents *Toxoplasma* antigens more efficiently, leading to CD8+ T cell activation (**Figure 4.C.i-ii.**) and parasite clearance. HLA polymorphisms are known to affect antigen-binding capacity. Sequence alignment with HLAs having available PDB structure showed A*33 shares 95.1% similarity with A*11 in global alignment (**Supplementary** Figure 2**.A., Table 2**), more than with A*24. The predicted structures confirmed greater 3D structural overlap of A*33 with A*11, particularly within the antigen binding pocket (residues 86–107), showing 86.3% similarity vs. 72.7% with A*24 (**Figure 5.A.i–ii.**). Ramachandran analysis validated stereochemical structure of A*33 model generated with A*11 template, with 83.7% residues in core and 7.2% in allowed regions (**Supplementary** Figure 2**.B.**). HADDOCK-based docking of A*33 and A*24 structures with 15 selected *Toxoplasma* peptides (**Figure 5.C., Table 3**) identified PLA2 (−98.7 ± 4.7 kcal/mol) and SAG2C (−104.2 ± 6.5 kcal/mol) as top candidates (**Figure 5.D.i–ii., iii-iv.**), due to stronger binding with A*33 and known immunogenicity in clinical infections[39],[40],[41]. Prior studies show that even single-point mutations in peptide can alter their HLA specificity[42],[43]. Amino acids residues in PLA2 and SAG2C interacting with antigen binding pocket of A*33 (**Table 4**) were subjected to unbiased in silico single-point substitutions with all 19 alternatives. Mutating key Ser57 in PLA2 yielded affinities similar to wild-type, suggesting stabilization through neighbouring interactions, making PLA2 more mutation-resilient. In contrast, replacing SAG2C Ser13 with Thr (S13T) disrupted binding with A*33 (−69.7 ± 2.3 kcal/mol), as did Arg21→Gln (R21E: - 75.4 ± 3.4 kcal/mol). Combining both mutations (S13T + R21Q) showed no further drop in affinity (−76.7 ± 5.1 kcal/mol) (**Figure 5.E.i–iii., Table 5**) confirming Ser13 as critical residue for SAG2C-A*33 stability. Eukaryotic expression and purification of recombinant A*33 and A*24 from K562 showed monodisperse SEC peaks (**Figure 6.A.i–ii.**), further confirmed by SDS-PAGE and anti-His western blot (∼42 kDa) (**Figure 6.B.i–ii.**). SPR analysis with immobilized A*33 and WT SAG2C peptide (analyte) showed a concentration-dependent binding response peaking at ∼150 RU at 200 µM, while A*24 showed minimal response (∼15 RU) at the same concentration (**Figure 6.C.i-iii.**). Further, S13T-mutant SAG2C peptide showed markedly reduced binding with A*33 (**Figure 6.C.iv.**), confirming that the structural features of A*33’s antigen-binding pocket enable specific interaction with SAG2C.

**Figure 5.**
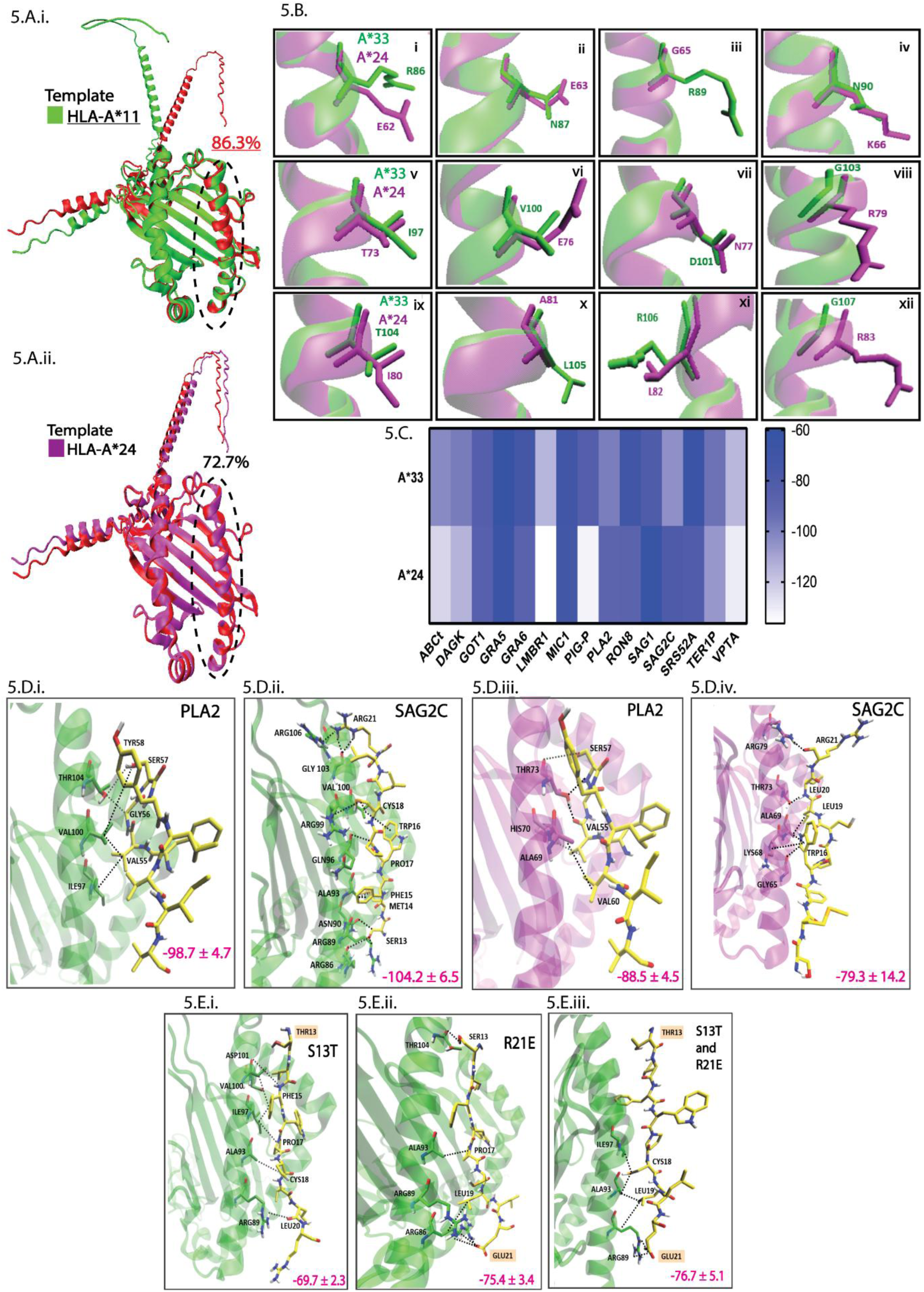
High affinity binding of HLA-A*33 to *Toxoplasma* antigenic peptides SAG2C and PLA2. **(A.i-ii.)** Superimposition of binding sites in A\**33 (*AlphaFold2 predicted) modelled structures using A*11 (green) and A*24 (purple) as template, respectively, with A*33 modelled without a template as the reference (red). Dotted lines highlight the binding site (residues 86–107), with A*11 showing the maximum similarity in binding site residues (86.3%)*. (**B.)*** Superimposed structures of variations in the binding site amino acids A*33 (green) (residues 86–107) and A*\*2*4 (purple) (residues 62–83) in panels (i–xii). (**C.**) Heatmap showing the differential binding HADDOCK score of 15 *Toxoplasma* antigenic peptides with A*33 and A*24. (**D.i-iv.**) Representative images of the interactions (black dotted lines) docked poses of *Toxoplasma* peptides PLA2 and SAG2C with A*33 and A*24, respectively. A*33 is depicted in green color, while A*24 is shown in purple color. Binding affinity is depicted pink. (**E.i-iii.**) Representative images of the interactions (black dotted lines) docked poses of the mutated *Toxoplasma* peptide SAG2C with A*33 (i.) Ser13 to Thr13 [S13T], (ii.) Arg21 to Glu21. [R21E], (iii.) Ser13 to Thr13 [S13T] and Arg21 to Glu21 [R21E]. Binding affinity is depicted pink.

**Figure 6.**
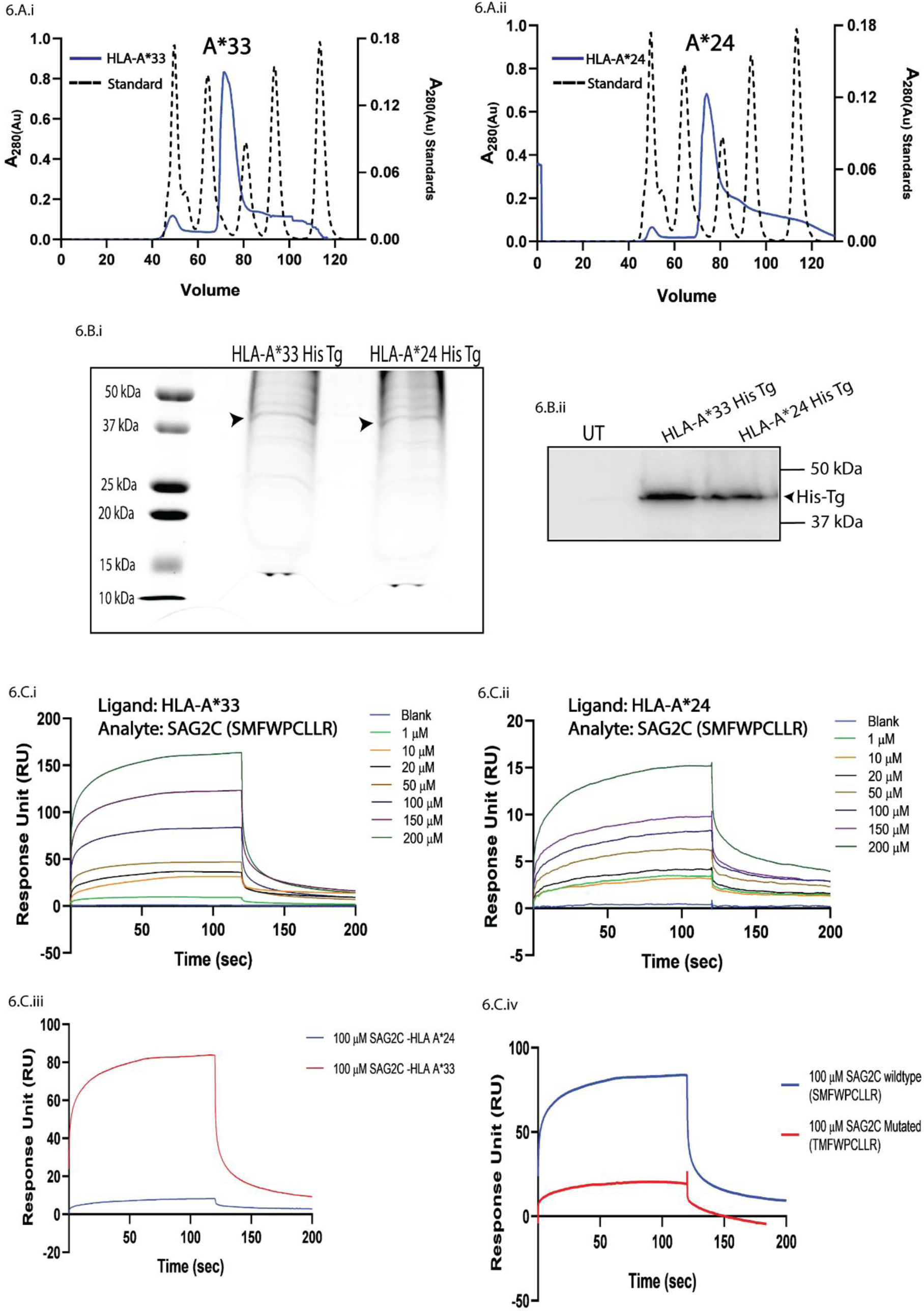
SPR analysis shows differential interactions between HLA-A*33 and HLA-A*24 with SAG2C peptide. (**A.i.-ii.**) SEC profile of purified A*33-His Tg (elution volume-71.2 mL) and A*24-His Tg (elution volume-74 mL) in Superdex 200 column, showed that it elutes as a monomer, respectively. The black dotted line shows the molecular weight standards. (**B.i.**) SDS-PAGE gel image of purified A*33-His Tg [∼42 kDa] and A*24-His Tg [∼42 kDa] is shown with the molecular weight standards (kDa) labelled on the left lane. (**B.ii.**) Western blot of whole cell lysate showing high expression of A*33-His Tg and A*24-His Tg (anti-His [∼42 kDa]) in transfected K562 cells, respectively, while no visible expression was observed in Untransfected (UT) cells, 168h post transfection. (**C.i.**) Concentration-dependent significant increase in response (RU) [∼150 RU] indicating strong binding, was observed when different concentrations of SAG2C (in μM) were injected as analyte over A*33 immobilized surface. (**C.ii.**) Concentration-dependent minimal increase in response (RU) [∼15 RU]) indicating relatively weaker binding, was observed when different concentrations of SAG2C (in μM) were injected as analyte over A*24 immobilized CM5 sensor chip surface. (**C.iii.**) Distinct decrease in SPR binding response was observed when similar concentration (100 μM) of SAG2C peptide was injected on the A*33 and A*24 immobilized CM5 sensor chip surface. (**C.iv.**) A similar sharp decrease in SPR binding response compared to wild-type SAG2C (100 μM) was observed when SAG2C mutant (100 μM) was injected on the A*33 immobilized CM5 sensor chip surface.

**Table 2:** Sequence similarity and root mean square deviation (RMSD) of non-hydrogen atoms for different A*\**33 models.

| Predicted Model | Template | Sequence Similarity (%) | Binding Site Similarity (%) | RMSD (Å) |
| --- | --- | --- | --- | --- |
| Model 1 | HLA-A*24 (PDB ID-7JYV) | 93.2 | 72.7 | 0.020 |
| Model 2 | HLA-A*11 (PDB ID-7S8Q) | 95.1 | 86.3 | 0.140 |

**Table 3:**
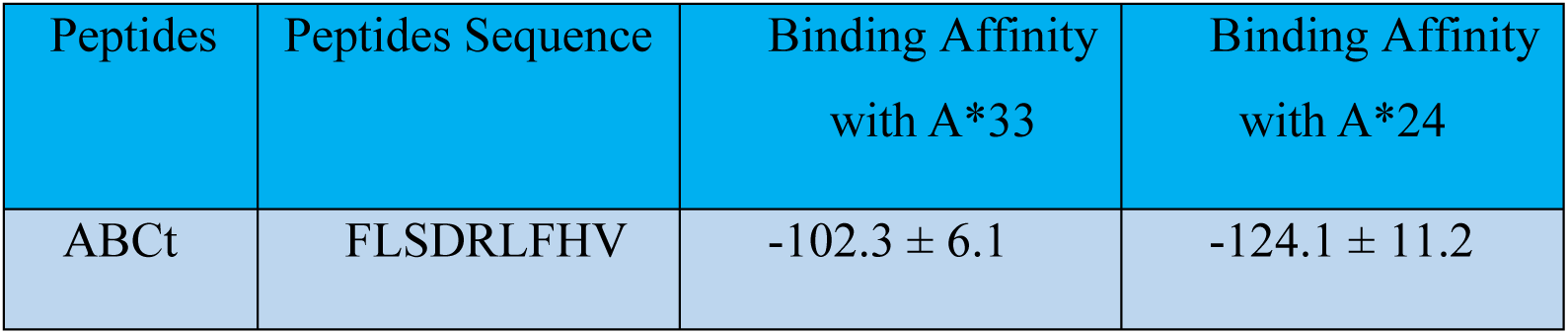
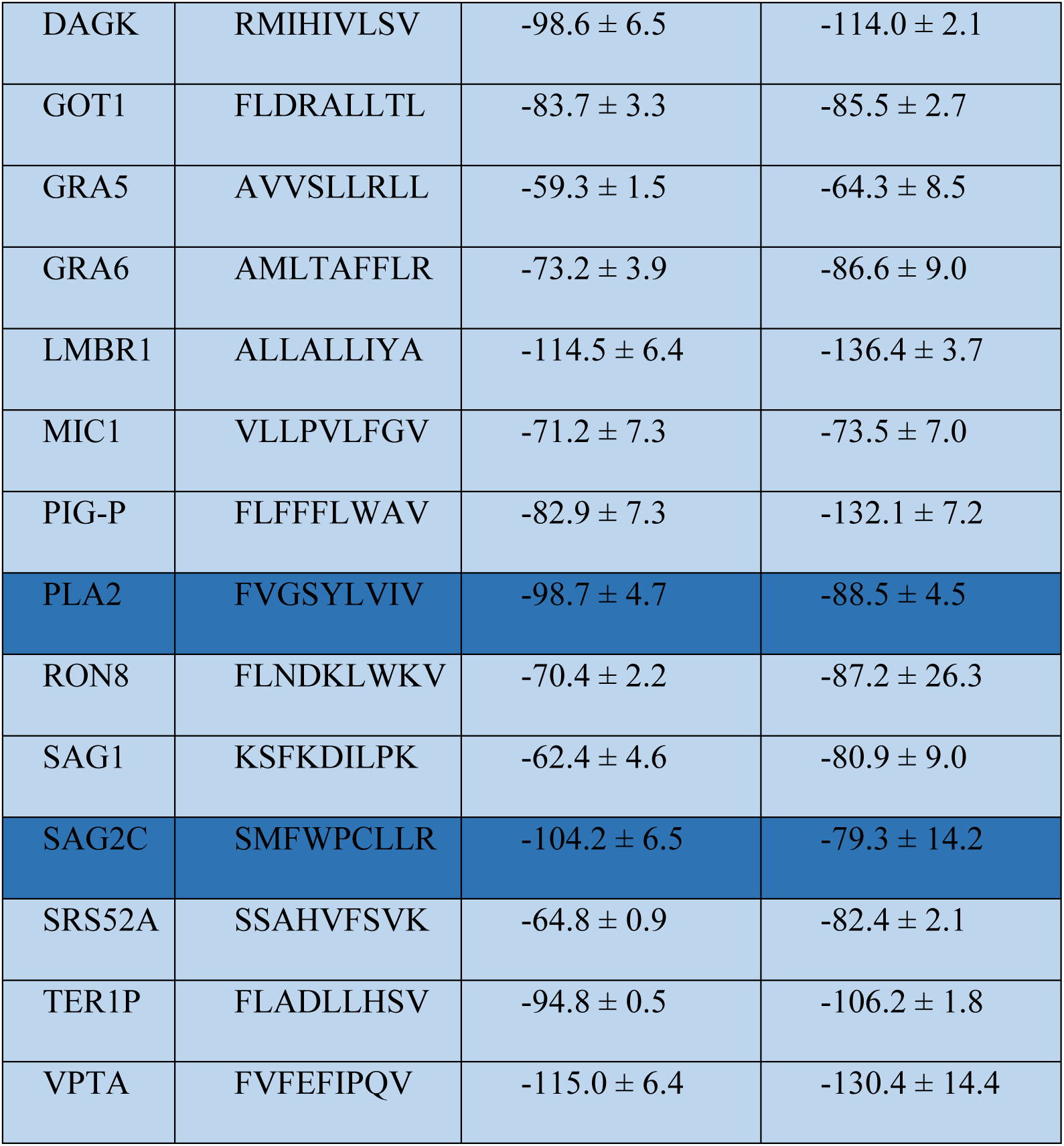
Peptide sequence and binding affinity values (kcal/mol) as computed from HADDOCK for the 15 peptides docked with A*33 and A*24 alleles. The standard error in the binding affinity values is also indicated.

**Table 4:**
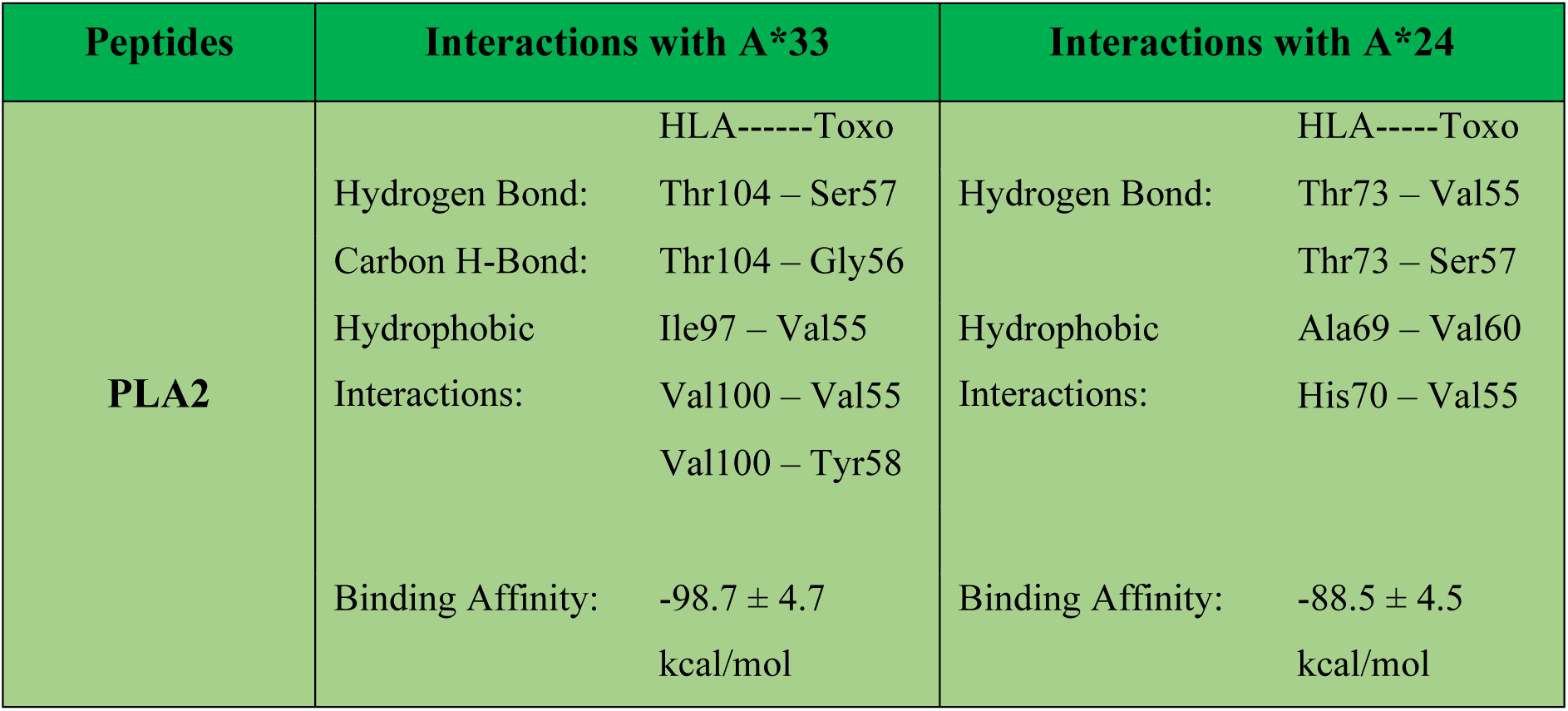
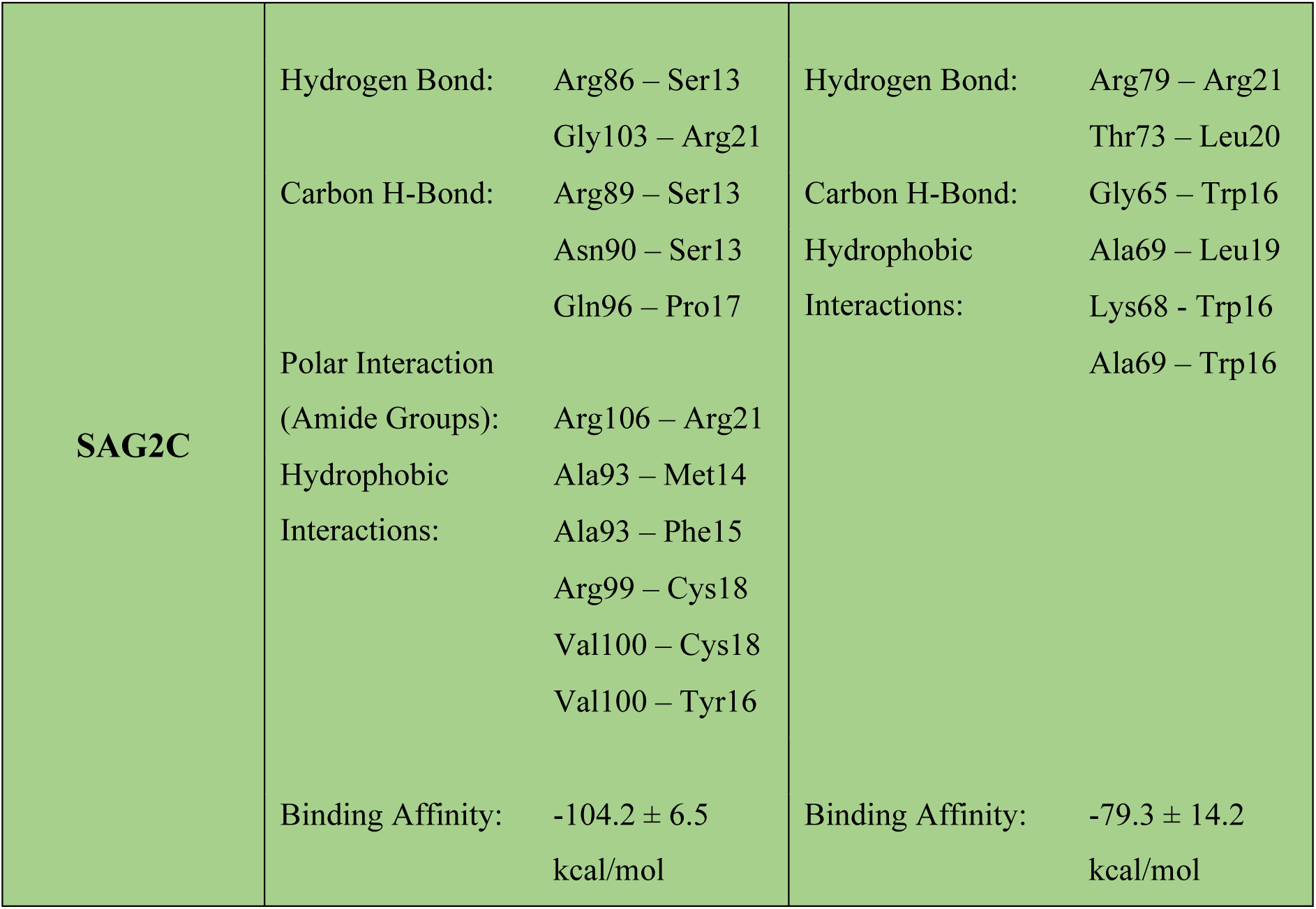
List of Residue-wise interactions of the selected peptide with A*33 and A*24.

| Peptides | Interactions with A*33 | Interactions with A*24 |
| --- | --- | --- |
| PLA2 | HLA-----Toxo | HLA-----Toxo |
|  | Hydrogen Bond: Thr104 – Ser57 | Hydrogen Bond: Thr73 – Val55 |
|  | Carbon H-Bond: Thr104 – Gly56 | Thr73 – Ser57 |
|  | Hydrophobic Ile97 – Val55 | Hydrophobic Ala69 – Val60 |
|  | Interactions: Val100 – Val55<br>Val100 – Tyr58 | Interactions: His70 – Val55 |
| | Binding Affinity: $-98.7 \pm 4.7$<br>kcal/mol | Binding Affinity: $-88.5 \pm 4.5$<br>kcal/mol |

|  |  |  |  |  |
| --- | --- | --- | --- | --- |
| **SAG2C** | Hydrogen Bond: | Arg86 – Ser13 | Hydrogen Bond: | Arg79 – Arg21 |
|  |  | Gly103 – Arg21 |  | Thr73 – Leu20 |
|  | Carbon H-Bond: | Arg89 – Ser13 | Carbon H-Bond: | Gly65 – Trp16 |
|  |  | Asn90 – Ser13 | Hydrophobic | Ala69 – Leu19 |
|  |  | Gln96 – Pro17 | Interactions: | Lys68 – Trp16 |
|  | Polar Interaction |  |  | Ala69 – Trp16 |
|  | (Amide Groups): | Arg106 – Arg21 |  |  |
|  | Hydrophobic | Ala93 – Met14 |  |  |
|  | Interactions: | Ala93 – Phe15 |  |  |
|  |  | Arg99 – Cys18 |  |  |
|  |  | Val100 – Cys18 |  |  |
|  |  | Val100 – Tyr16 |  |  |
|  | Binding Affinity: | -104.2 ± 6.5 | Binding Affinity: | -79.3 ± 14.2 |
|  |  | kcal/mol |  | kcal/mol |

**Table 5:** The binding affinities (in kcal/mol) for mutated PLA2 and SAG2C peptides with A*33. Highlighted scores indicate mutations with lower affinity, showing better interaction with the wild type compared to the mutated sequence. (The binding affinity values are in kcal/mol)

| Mutated<br>Amino Acid | PLA2 | SAG2C |  |
| --- | --- | --- | --- |
|  | Position: Serine 57<br>(-98.7 ± 4.7) | Position: Serine13<br>(-104.2 ± 6.5) | Position: Arginine21<br>(-104.2 ± 6.5) |
| Ala | -96.7 ± 1.2 | -88.0 ± 1.9 | -84.5 ± 3.5 |
| Arg | -101.0 ± 19.0 | -86.6 ± 2.7 | - |
| Asn | -111.9 ± 6.5 | -75.3 ± 0.9 | -84.4 ± 3.0 |
| Asp | -111.6 ± 3.3 | -92.3 ± 4.2 | -84.6 ± 4.0 |
| Cys | -96.3 ± 5.0 | -89.9 ± 5.2 | -77.2 ± 0.6 |
| Gln | -98.8 ± 6.9 | -84.1 ± 2.1 | -76.6 ± 3.2 |
| **Glu** | -110.3 ± 5.4 | -82.1 ± 3.6 | **-75.4 ± 3.4** |
| Gly | -97.2 ± 2.4 | -107.7 ± 7.6 | -91.8 ± 4.2 |
| His | -105.3 ± 3.9 | -110.9 ± 1.7 | -87.7 ± 1.0 |
| Ile | -107.6 ± 3.6 | -76.2 ± 1.7 | -89.9 ± 2.3 |
| Leu | -109.3 ± 4.8 | -105.9 ± 6.4 | -88.2 ± 4.2 |
| Lys | -107.2 ± 6.3 | -85.0 ± 1.5 | -107.0 ± 6.2 |
| Met | -105.9 ± 2.7 | -88.7 ± 5.1 | -106.1 ± 2.6 |
| Phe | -107.2 ± 10.2 | -113.2 ± 5.0 | -97.3 ± 6.7 |
| Pro | -101.6 ± 3.5 | -81.7 ± 1.2 | -88.6 ± 5.0 |
| Ser | - | - | -79.2 ± 2.5 |
| **Thr** | -107.5 ± 7.6 | **-69.7 ± 2.3** | -103.2 ± 2.3 |
| Trp | -112.8 ± 6.4 | -88.5 ± 2.0 | -102.7 ± 2.1 |
| Tyr | -92.6 ± 3.5 | -104.0 ± 2.6 | -95.7 ± 4.0 |
| Val | -111.4 ± 3.7 | -88.2 ± 2.1 | -100.0 ± 1.0 |

### Ectopically expressed HLA-C*07-K562 restricts *Plasmodium* proliferation

Since A*33 displayed a protective role against *Toxoplasma*, the role of C*07 previously identified to negatively correlate with *Plasmodium* infection (**Figure 1.F.ii.**) was further evaluated to determine if such protective role is restricted only to A*33 or could be attributed as a generalized mechanism in context of other Apicomplexan infection. C*07 was expressed in HLA class-I-null K562-cells[44] (**Figure 7.A.**) and confirmed by flow cytometry (**Figure 7.B.i–iii.**). For *Plasmodium* infection model, C*07-K562 cells were erythroid-differentiated using EPO[44],[45],[46],[47], showing robust CD235a+ (glycophorin-A) expression (**Figure 7.A., 7.C.i–ii.**), allowing in vitro infection assays, with previously reported *Plasmodium* susceptible allele, A*33-K562[48] as control (**Supplementary** Figure 1**.A–C.**), although our NGS revealed no significant association of the same, we still proceeded to check A*33 as a susceptible experimental control. While early-stage parasites were observed in both C*07 and A*33 expressing K562 at 6h P.I., with comparable ring-stage parasite at 12h P.I. (**Figure 7.D.i–ii.**), major differences emerged as infection progress. While C*07-K562-cells halted parasite development at the ring stage, blocking progression to trophozoite/schizont stages and clearing the infection by 48h (**Figure 7.D.iii.**), A*33-K562-cells, in contrast, supported continued parasite maturation, leading to host cell damage and nuclear fragmentation at 48h P.I. (**Figure 7.D.iii.**). Subsequently, the infectivity index was significantly higher in A*33-expressing cells, while C*07-expressing cells effectively restricted parasite growth (**Figure 7.D.iv.**). These findings suggest that C*07 confers protection against *Plasmodium* infection by impairing progression to the mature stages, highlighting HLA specificity as a key factor in determining outcome of Apicomplexan infection.

**Figure 7.**
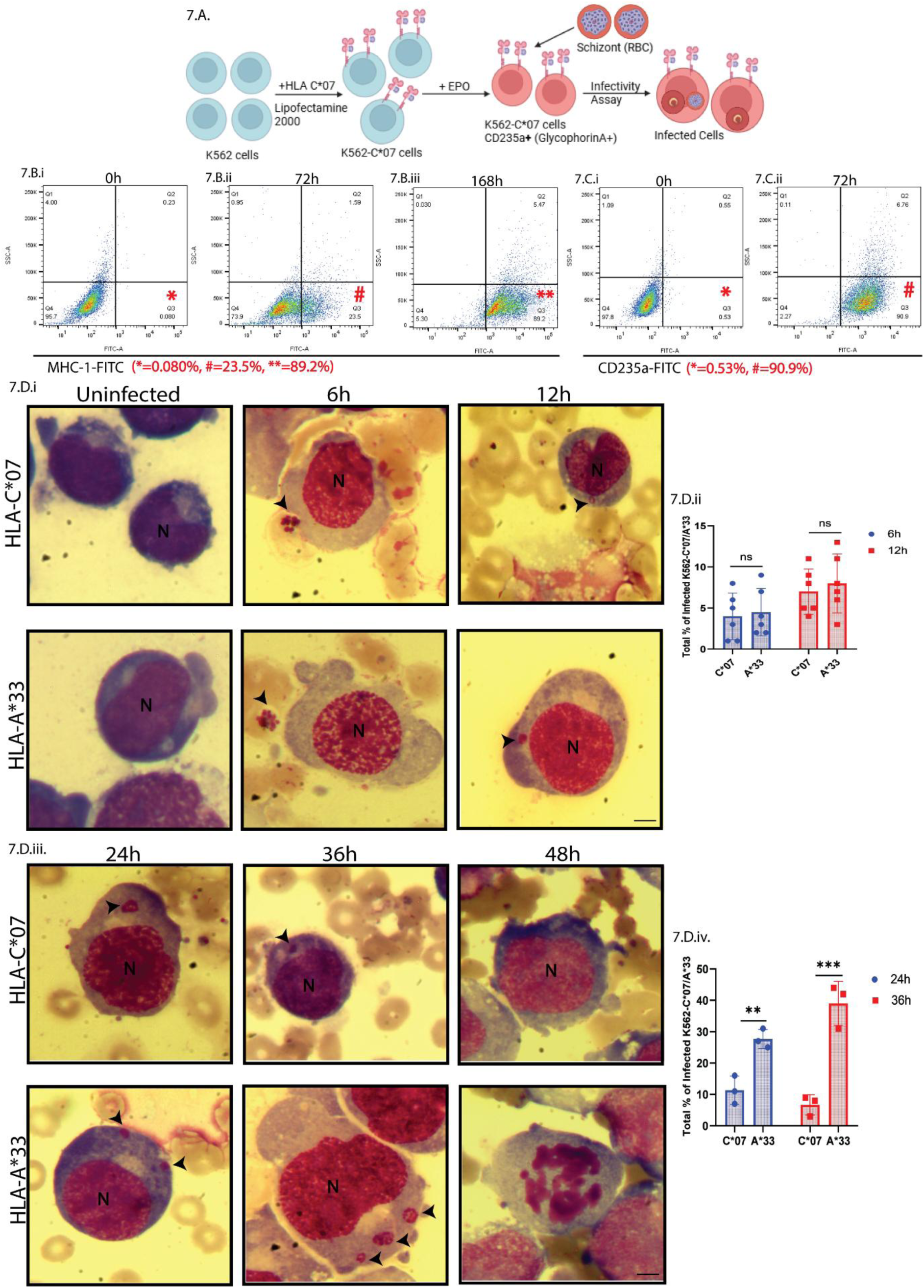
C*07 stymies successful expansion of ring stage *Plasmodium* in spite of similar infectivity in early stages of infection. **(A.)** Schematic representation showing *Plasmodium* infection in C*07 transfected-erythroid differentiated K562 cells. **(B.i.-iii**.) Representative Flow cytometry-based scatter plot showing significant proliferation in population of FITC-MHC-I cells, 72h and 168h post C*07 transfection (T) in K652 cells. (**C.i.-ii.**) FACS plot showing different proportions of FITC-CD235a+ erythroid cells 72h post EPO treatment in K562 cells. (**D.i.**) Representative Giemsa stain images of C*07 (T) and A*33 (T) infected cells at 6h and 12h P.I. Black arrow represents *Plasmodium* parasites. N denotes host nucleus. Scale bar 5μM. (**D.ii.**) Bar graph plot of Infectivity Index for 6h and 12h P.I. Each dot represents count from 100 infected C*07 (T) and A*33 (T) cells (n=6). (**D.iii.**) Representative Giemsa stain images of C*07 (T) and A*33 (T) infected cells at 24h, 36h and 48h P.I. Black arrow represents *Plasmodium* parasites. N denotes host nucleus. Scale bar 5μM. (**D.iv.**) Bar graph plot of Infectivity Index for 24h and 36h P.I. Each dot represents count from 100 infected C*07 (T) and A*33 (T) cells (n=6). Data is represented as Mean ± SD from 3 independent experiments. p ≤ 0.05 is marked as *, p ≤ 0.01 is marked as ** and p ≤ 0.001 is marked as ***.

## Discussion

The management of HbE/β-thalassemia, a disorder arising from compound heterozygosity of the HbE mutation and β-thalassemia, is particularly challenging due to its pronounced clinical heterogeneity[2]. Accounting for approximately 15–20% of severe thalassemia cases, the disease is marked by asymptomatic to severe anemia, repeated blood transfusion dependence, and other co-morbid conditions, making it very challenging for clinicians to manage the disease. While the core genetic mutations are well-documented, phenotype-modifying factors such as infections and immune components have largely been overlooked although they are critical factors in defining disease severity. In this regard, a higher proportion of HbE/β-thalassemia patients from regions of co-endemicity exhibit have been shown to exhibit *Plasmodium* positivity[49],[50], and prior studies suggest that endemic protozoan infections like *Plasmodium* and *Toxoplasma* exert selective pressures influencing the disease’s clinical[51]. Interestingly, in this study, initial testing for infection positivity (*Plasmodium* and *Toxoplasma*) among a cohort of HbE/β-thalassemia patients from Eastern India revealed no clear correlation with the frequency of blood transfusions. Paradoxically, increased transfusion frequency was associated with a decline in infection positivity **(Figure 1**.), despite patients being asymptomatic and transfusion protocols being strictly followed. This indicates that transfusion-related infection risk[52],[53] may not solely depend on frequency, challenging earlier assumptions[54],[55]. Moreover, Next-Generation-Sequencing data of individual HbE/β-thalassemia patient samples, indicated protective effects against common Apicomplexan *Plasmodium* and *Toxoplasma* infections, among HbE/β-thalassemia patients to be linked with genetic architecture of their Human Leukocyte Antigen (HLA) C*07, DQB1*03, and A*33. In contrast, increased susceptibility to *Plasmodium* infections was found to be associated with the presence of the DQB1*02 allele in these patients.

Given the role of HLAs in immune surveillance, their high diversity has been linked with evolutionary adaptations to a broad range of pathogens[56],[57]. Importantly, HLA polymorphisms have been implicated in parasitic infection susceptibility[58] and thalassemia severity[10],[59].

Ex vivo infection assays validated these associations. PBMCs carrying the A*33 allele, which correlated negatively with *Toxoplasma* infection, allowed parasite invasion but effectively inhibited intracellular proliferation, leading to clearance, unlike A*24 PBMCs, which showed higher parasite load leading to host cell lysis (**Figure 2 and 3**.). Mechanistic studies further demonstrated upregulated HLA-I expression (**Figure 4**.) and increased IFN-γ-producing CD8+ T cell subsets in A*33 infected PBMCs as compared to A*24 (**Figure 4**.), suggesting enhanced antigen presentation and immune activation.

Despite minor differences in HLA primary sequences, even single nucleotide variations in the antigen-binding pocket can impact immune responses[60]. Structural flexibility of MHC (HLA) has been previously identified as critical factor, making host cells either permissible or non-permissible to protozoan infection[61],[56]. Our study highlights *Toxoplasma* antigenic peptides SAG2C and PLA2, which are dominantly represented in clinical infection[62],[63], bind more strongly to A*33 than to A*24 (**Figure 5**.). SPR analysis confirmed higher affinity of SAG2C to recombinant A*33, with this interaction disrupted upon altering key binding residues (**Figure 6**.).

Importantly, A*33-mediated protection appears specific to *Toxoplasma*, as *Plasmodium falciparum* infection studies implicated C*07, not A*33, in protective responses (**Figure 7**.). While A*33 has been recently associated with *Plasmodium* susceptibility in certain populations[48], our NGS data showed no significant association of A*33 within HbE/β-thalassemia cohort of Eastern India, though experimental data suggested mild susceptibility. This discrepancy may reflect complex interactions between HbE and infection susceptibility, possibly contributing to balancing selection and regional HLA enrichment.

Although this is an eastern India focused study, correlation from multiple publicly available databases, like IndiGEN [64], highlight the lower prevalence of A*33, in areas where both HbE/β-thalassemia trait [2] and *Toxoplasma* infection is significantly low (8.8%), like that in the western part of India [65].

These allele-pathogen dynamics underscore the selective fitness advantage conferred by specific HLA variants in the context of HbE/β-thalassemia as summarised schematically in the graphical abstract. Such insights are critical for understanding HLA distribution in endemic regions like India, which bears a disproportionate burden of neglected tropical diseases, accounting for 83% of malaria cases and 82% of malaria deaths in South-East Asia. The findings advocate for personalized clinical management strategies and highlight the evolutionary implications of allele perpetuation in affected populations[56].

## Ethical Statement

The purpose of the study was explained to participants and written informed consent was obtained prior to the study. The ethical approval for the study was obtained from both the Institute as well as NRS Medical College prior to collection. (Approval Nos. IIT/SRIC/DR/2019 dated 06.11.2019, IIT/SRIC/DEAN/2022 dated 12.08.2022; No/NMC/3483 dated 12.07.2019; IIT/SRIC/SAO/2017 dated 11.12.2017; No/NMC/4154 dated 03.08.2016).

## Data Sharing Statement

The sequencing data are not publicly available due to privacy or ethical restrictions. It has already been deposited in the SRA database (NCBI) with BioProject accession number PRJNA1251014 (https://www.ncbi.nlm.nih.gov/sra).

## Data Availability

All data produced in the present work are contained in the manuscript

## Acknowledgements

Authors would like to acknowledge Confocal Microscope facility of BSBT, DST-FIST, Govt. of India, FACS facility of BSBT (Dr. Ritobrata Goswami) and CRF-SMST, IIT KGP (Dr. Gayatri Mukherjee), SPR facility of CRF-IIT KGP (Dr. Dibyendu Samanta). Authors would like thank all the blood donors. Authors would like to acknowledge Professor Swati Patankar, IIT Bombay; Dr. Abhijit Deshmukh, NIAB, Hyderabad and Professor Dhanasekaran Shanmugam, CSIR-NCL, Pune for their kind help in procuring *Toxoplasma gondii* (ΔKu80) and GFP-ΔKu80 parasites and human foreskin fibroblasts (HFF) cells. SAG-1 and GAP45 antibodies and 2.8 lic HisTag plasmid were a kind gift from Prof. Dominique Soldati-Favre, Department of Microbiology and Molecular Medicine, University of Geneva.

## Authorship Contribution

SB conceptualization, sample collection, experiments, analysis, MS preparation. MR sample collection, analysis, MS preparation. SS helped with bioinformatics analysis and modelling studies. DKR helped in performing experiments related to *Plasmodium spp*. SP and BD conceptualization and clinical insights. PCS and GM helped in conceptualization and project administration. MG did bioinformatics analysis, MS review and editing. MKB helped with *Plasmodium* experiments, MS review and editing. TKD helped in sample collection, conceptualization, clinical insights and MS review. NC and BM conceptualization, conceived and directed the project and prepared the MS with input from all the authors. All authors have read and approved the final version of the manuscript.

## Supplementary File

## Supplementary Methodology

### Sample Collection and Classification of the HbE/β-thalassemia patients based on disease severity

To study the phenotypic heterogeneity of the HbE/β-thalassemia, patients were stratified into three different disease severity groups (mild, moderate and severe) as per Mahidol Scoring (ref), a standard scoring method used to grade clinical severity of HbE/β-thalassemia patients. Peripheral blood (2-4ml) was collected from each participant in EDTA tubes. Demographic parameters of the enrolled patients were evaluated.

### HLA haplotying of the patients using high-resolution NGS and Sanger sequencing method

For Illumina based NGS sequencing, long range PCR primers were employed with genomic DNA to amplify the individual HLA loci. Genomic DNA was fragmented and ligated with sequencing adapters to produce a library that was sequenced on Illumina HiSeq sequencer to generate 2x100 bp sequence reads. The generated sequence data was analysed after necessary quality control for variant calling and annotation. The raw sequence reads are aligned using a FASTA-like algorithm. All data were then analysed to infer any possible corelation and association between HLA alleles and disease phenotype. Statistical tests were run to find the association with specific severity groups. All samples were screened for HbE/β-thalassemia and β-globin mutation using ARMS-PCR followed by Sanger sequencing as already reported in (MR Gene reports paper).

### qRT-PCR (Real-time quantitative polymerase chain reaction) and High-Resolution Melting Curve Analysis (HRM) to detect the presence of *Plasmodium* and *Toxoplasma* infection in HbE/β-thalassemia patient blood

Each reaction was carried out in a total volume of 10ul containing 5ul of PowerUp^TM^ SYBR Green Master Mix (2X), 1ul of each of reported primers for specific detection of *Plasmodium sp* and *Toxoplasma*. (**Table 1)**, 1ul of each clinical DNA sample and rest volume made up to 10ul using Nuclease-Free water. DNA from parasites (*Plasmodium sp* and *Toxoplasma)* and healthy individuals was used as control in each case. MicroAmp^®^ Fast Optical 0.1 ml 96-well reaction plate (Applied Biosystems), sealed with MicroAmp^®^ Optical adhesive film (Applied Biosystems), were used to perform qRT-PCR-HRM. The tubes or plates were centrifuged briefly prior to undergoing qRT-PCR-HRM. The qPCR products were subject to a melt program: denaturation of double stranded DNA at 95°C for 15 seconds, annealing of double stranded DNA at 60°C for 1 minute, followed by a gradual temperature increase(dissociation) until 95°C for 15 seconds (annealing). The data analysis was carried out using the ΔΔCT method. Annealing temperature was at 59^0^ C. All Real-time PCR reactions for HLA detection were also performed using QuantStudio 5 real-tme PCR systems (Applied Biosystems™). All reactions were performed in at least three different samples in triplicate.

### Peripheral Blood Mononuclear Cell (PBMC) Isolation and Maintenance

Blood was collected from both HLA-A*33+ (referred as A*33 in subsequent text) and HLA-A*33-(HLA-A*24+ (Table 2) (common allele from positive patients as per NGS)) (referred as A*24 in subsequent text) individuals in EDTA coated vials and slowly inverted to mix blood gently for 10-15 mins and the blood was transferred in a 50ml falcon tube. 1X PBS BUFFER was used for diluting blood in ratio 1:6 and mixed well. Ficoll-paque was taken in a 15ml falcon tube in 1:1 ratio to the blood collected. The falcon tube was is then kept in a tilted position and the PBS-BLOOD mix was slowly layered over the ficoll through the wall of the falcon tube (the layer should be undisturbed). Then it was centrifuged at 400g for 50 minutes with no brakes at 18°-20°C. Plasma layer was removed and the MNCs were transferred in a fresh 15ml falcon tube. 10ml 1X PBS buffer was added for washing the cells and centrifuged at 300g for 30 minutes at 20°C. Washing was repeated twice using 1X PBS at 200g for 30 minutes, to remove any residual platelets. Cell pellet is suspended in 300ul complete RPMI media and plated in 12 well plate and incubated in CO_2_ incubator. After 24 hours GMCSF supplement was given to the cells.

### Immunofluorescence Assay (IFA) and Confocal Microscopy

Initially blocking was done using 250ul/well 2% BSA/PBS and was kept in shaker at 4°C. Subsequently, cells were stained with anti-*Toxoplasma* surface protein SAG1 (1:3000), without permeabilization to discriminate between intracellular and extracellular parasites, and incubated for 30 minutes in 2% BSA/PBS in shaker at 4°C. Then the wells are washed four times, each wash for 5 minutes in 2% BSA/PBS. Further, the permeabilization was performed by adding 0.2% triton in PBS and incubated in shaker for 20 minutes. Again, the blocking was performed by adding 2% BSA, 0.2% triton in PBS solution to the wells and incubating for 20 minutes in the shaker. Anti GAP45 (1:5000) in 2% BSA, 0.2% triton in PBS solution was added to the wells and incubated in shaker for 1hr. After the incubation, washing was performed 3 times, each wash is for 5 minutes, using 0.2% triton in PBS. Cells were then incubated with secondary antibodies (Alexa488-or Alexa594-conjugated goat anti-mouse or goat anti-rabbit IgGs) for 1hr. Washing was performed again, 5 minutes each using 0.2% triton in PBS. Parasite and PBMC nuclei were stained with DAPI (40,6-diamidino-2-phenylindole; 50µg/ml in PBS), and coverslips were mounted on Fluoromount G (ThermoFisher Scientific Cat. No. 00-4958-02) on glass slides and stored at 4°C in the dark. mCherry-HLA transfected K652 cells were also assessed for the tachyzoite growth kinetics by infecting with the GFP-ΔKu80. Cells were fixed at different time points to study the infectivity index. DAPI (TC229) was purchased from Himedia. Fluoromount-G (00-4958-02) was obtained from Invitrogen. All images were captured using Olympus confocal microscopy (FV3000) and Leica Stellaris 5 Dmi8 demonstration unit. Final image analysis and processing were done with ImageJ.

### Apoptosis Assay

For the apoptosis assay, PBMCs (A*33+, A*33-) were seeded at a density of 0.1 million cells per well onto 12-well flat-bottom tissue culture plates and infected with freshly egressed tachyzoites. Cells were then cultured in a CO2 incubator at 37°C, and harvested at different time points, and rinsed with PBS. Infection mediated apoptosis was detected by Annexin V Conjugates for Apoptosis Detection staining (Invitrogen), where the binding of FITC-conjugated Annexin V to the apoptotic cells was analyzed using flow cytometry.

### Cloning and over-expression of mCherry-A*33 and mCherry-A*24 alleles

The full length (768bp) of mCherry was amplified using primers (forward: 5’-ctggctagcgtttaaacttaagcttATGGTGAGCAAGGGCGAG-3’, reverse:5’-ggggcgccatgacggccatggatccCTTGTACAGCTCGTCCATGC-3’) from pcDNA3.1-mcherry (a gift from David Bartel, Addgene plasmid # 128744). The PCR product was then ligated with the BamHI-HF and HindIII-HF restriction enzyme (NEB) digested A*33/A*24-pcDNA3.1 backbone plasmids (A*33:01:01 was a gift from Derin Keskin & Catherine Wu (Addgene plasmid # 203288) (A*24:02:01 was a gift from Derin Keskin & Catherine Wu (Addgene plasmid # 203295) using the NEB Gibson Assembly Master Mix (Lot #10229799). Post transformation, positive colonies were screened and confirmed via restriction digestion (BamHI-HF, NotI-HF and HindIII-HF restriction enzymes) and PCR amplification. Thereafter, pcDNA3.1-mcherry/HLA construct was transfected into K562 cells (HLA null cells) using Lipofectamine™ 2000(Invitrogen) and screened with 1000µg/ml G418 disulfide (A1720, Sigma) for over 7 days. Transfection with pcDNA3.1-mcherry empty vector (EV) was acted as control. The positive clones were confirmed by RT-qPCR, IFA and Western Blot for mcherry-HLA-33/24 expression. The primers for mCherry A*33 were as follows: forward,5’-ACAACCAGAGCGAGGCCGGTG -3’; reverse,5’-CCCAAGGCTGCTGCCGGTGTG -3’. Annealing temperature was at 59^0^ C. The primers for mCherry HLA-A*24 was as follows: forward,5’-AACCCTCCTCCTGCTACTCTCG -3’; reverse,5’-TCTGGATGGTGTGAGAACCTGGC -3’. Annealing temperature was at 59^0^ C as well.

### Cloning and purification of C-terminal His-tg-A*33 and His-tg-A*24 proteins

The full length of 6X His-Tg was amplified using primers (forward,5’-ctctcacagcttgtaaagtgGTGCTGTTTCAGGGCCCC-3’; reverse,5’-ggatatctgcagaattctcaGGGGAGGTGTGGGAGGTT-3’) from the 2.8 lic HisTag plasmid. The PCR based linearization of the A*33 plasmid was done using primers (forward,5’-TGAGAATTCTGCAGATATC; reverse,5’-CACTTTACAAGCTGTGAG). The linearization of the HLA-A*24 plasmid was done using primers (forward,5’-TGAGAATTCTGCAGATATC; reverse,5’-CACTTTACAAGCTGTGAG). The PCR products were then ligated separately for the two alleles, using the NEB Gibson Assembly Master Mix (Lot #10229799). Post transformation, positive colonies were screened and confirmed via restriction digestion (BamHI-HF, NotI-HF and HindIII-HF restriction enzymes) and PCR amplification. Thereafter, His-Tg/HLA construct was transfected into K562 cells using Lipofectamine™ 2000(Invitrogen) and screened with 1000µg/ml G418 disulfide (A1720, Sigma) for over 7 days. Then they were used for protein purification, following harvesting and lysis of cells, as per previously used protocols (ref). The supernatant collected post centrifugation was subjected to nickel-nitrilotriacetic acid (Ni-NTA) based affinity chromatography. The eluted fractions were collected in different concentrations of Imidazole (ref) and subject to concentration using the 50 kDa Amicon ultra centrifugal filter cut off column (Merck). The protein was next subjected to SEC and the peak fractions were collected. The purified protein fractions were then run on a 12% SDS gel to assess their purity, and further protein expression was confirmed by immunoblotting using anti-HisTg antibody (His-Tag (D3I1O) XP Rabbit mAb #12698) directed against the HisTg protein.

### Immunoblotting analysis

Post transfection and drug treatments, K562 cells were scraped harvested by centrifugation and lysed in 2X SDS sample loading buffer consisting of 10% β-mercaptoethanol, pH=6.8. Samples were boiled at 95°C for 10 minutes and subjected to SDS-PAGE. Resolved proteins were then transferred onto a nitrocellulose membrane (BioRad) using a semi-dry transfer system (BioRad). Blocking of membranes were done in 5% milk in 0.05%Tween20/PBS for 45 minutes at room temperature followed by incubation in primary antibodies (anti-MHC-I Class I (EMR8-5) mouse mAb #88274, anti-mCherry rabbit polyclonal anti body, anti-His-Tag rabbit (D3I1O) XP) (Cell signalling technology) diluted in blocking solution overnight at 4°C in an orbital shaker. Membranes were then incubated with the HRP conjugated goat anti-rabbit IgG or goat anti-mouse IgG secondary antibody. SuperSignal West Pico PLUS Chemiluminescent Substrate (Thermo Scientific) was used as substrate solution and membranes were incubated in it for developing and the chemiluminescence blot images were taken in Chemidoc MP Imaging System (Bio-Rad). β-actin was used as a positive loading control for the whole cell lysate.

### Structural Modelling of A*33 and Peptide Selection for Docking Studies

Since MHC Class I (HLA I) Heavy Chain α covers the entire binding groove domain ^1^,^2^ and IMGT/HLA database primarily contains Chain A’s exact sequences, it is the only chain considered for the alignment and homology modelling, as previously reported^3,4,5^. The sequence for A*33 obtained from the IPD-IMGT/HLA Database^6^ (Accession No. HLA00104), was aligned with three HLA-A alleles: HLA-A*24 (PDB ID: 7JYV), HLA-A*29 (PDB ID: 7TLT), and HLA-A*11 (PDB ID: 7S8Q) using CLUSTAL Omega^7^. The structure of A*33 was modeled using AlphaFold2 on Google Colab, called Colabfold^8,9^ using the templates HLA-A*24, HLA-A*29, and HLA-A*11. The modeled structures of A*33 using different templates were aligned using VMD^10^ (Visual Molecular Dynamics). A*33 structure obtained without using any template was used as reference to compute the root mean square deviations (RMSD) of the non-hydrogen atoms of the alleles with the reference structure. A total of 16 peptides derived from the protozoan parasite *Toxoplasma* were chosen for this study. They were selected on the basis of a rational to select immunogenic peptides that have been previously reported (ref) to induce IFN-ℽ mediated response against Toxoplasmosis in human leukocytes and other animal models of the disease. The peptide sequences were obtained from ToxoDB^11^ (http://ToxoDB.org), as listed in **Table 5**. HLA-A*24, the most common allele among the *Toxoplasma* positive patients based on the NGS data, is used here as the control (susceptible) allele for the binding studies. The 15 peptides were docked with both the A*33 model and the A*24 template for comparative analysis using HADDOCK, which provides the HADDOCK scores to measure the binding affinity. HADDOCK was chosen in this study as it is widely recognized for its reliability in peptide-protein docking^12,13^. HADDOCK’s default parameters allowed flexible conformational adjustments to model realistic interactions. The binding interactions of 2 peptides (PLA2, SAG2C) with A*33 and A*24 were subsequently analysed using Discovery Studio 2024 (BIOVIA DS, San Diego: Dassault Systèmes, 2024) to identify the key residues involved in binding to assess the stability of the complexes. These key amino acids were mutated by the other 19 amino acids and modelled using Alphafold2. Consequently, both single-point and double mutations were performed for the peptides. These mutated peptides were again docked with the HLA-A*33 and A*24 using HADDOCK with the same protocol discussed above. These analyses offer valuable insights into the structural basis of peptide-HLA interactions and their influence on binding specificity and stability.

### Surface plasmon resonance for protein-peptide interaction study

The interaction of A*33, A*24 with wild type (WT) SAG2C and A*33 with mutant (MUT) SAG2C was analyzed using Biacore T200 at 25◦C. The A*33 protein was immobilized on a Series S CM5 chip (Cytiva) using amine coupling method to a final response of 200 RU (Response units) in both cases. SPR buffer (1 M HEPES (pH 7.4), 1 M NaCl, 0.5 M EDTA, 1ml Tween-20) was used as the running buffer throughout the experiment and different concentrations of SAG2C (WT, MUT) as an analyte (1–200 μM) were injected at a flow rate of 30 μl/min for 120s. The experiments were performed in duplicates and after each cycle of experiment, regeneration of the surface was done using 10 mM glycine, pH 2.5. The final response was obtained by subtracting the response of the blank channel (with no protein immobilized) to eliminate the effect of the non-specific interaction of analyte to the sensor chip. The results were analyzed using BIA evaluation Software 2.0 and Graph Pad Prism 8.0.

### Statistical Analysis

The statistical analysis was carried out using GraphPad Prism 8 software. The results were presented as Mean ±SD and one-way ANOVA was performed to compare the study groups. Descriptive statistics were carried out for demographic and hematological data. Fisher’s exact test was used in case of observed counts less than 20. Odds ratio (ORs) and 95% confidence intervals were calculated as statistical differences regarding HLA frequencies. p-value was corrected by Bonferroni corrections in multiple comparisons to eliminate Type І errors. p-values <0.05 were considered to be statistically significant. Individual data points representing biological replicates were included in the graph. All experiments were repeated at least three times and in triplicates. The comparison between two groups were performed by two-tailed Student’s t-test.

## Supplementary Figures

**Supplementary Figure 1.**
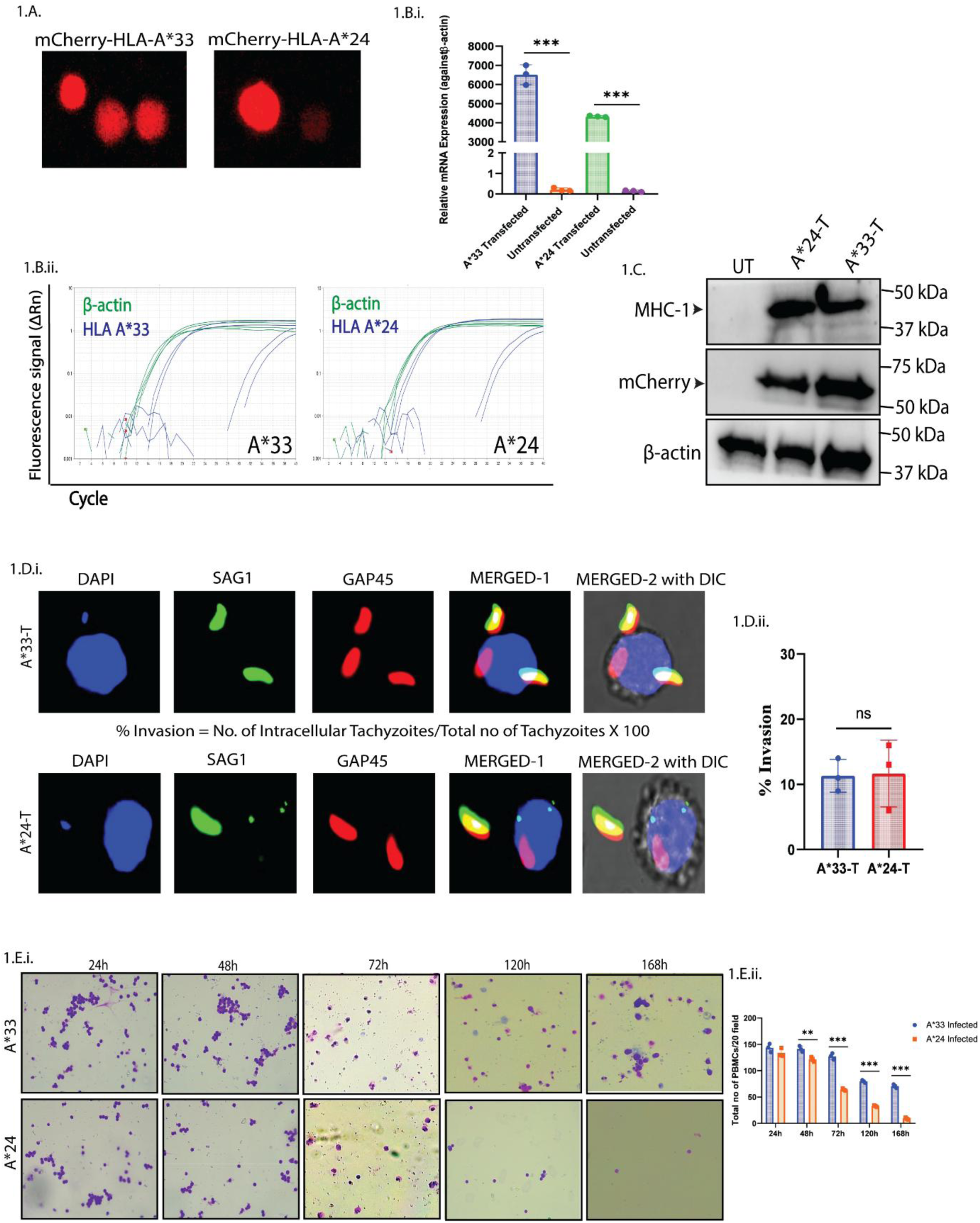
**(A.)** Representative Confocal images of the mCherry tagged A*33 and A*24 expression in the transfected K562 cells, 168h post transfection **(B.i.)** Relative A*33 and A*24 expression fold change (2^ΔΔCt) in the transfected K562 cells, 168h post transfection, was determined from qRT-PCR depicting the HLA expression in each group with respect to β-actin. (**B.ii.**) qRT-PCR based Amplification plot of A*33 and A*24 expression in the transfected K562 cells. (**C.**) Western blot of whole cell lysate showing high expression of mCherry-A*24 and mCherry-A*33 (anti-MHC-I [∼42 kDa] and anti-mCherry [∼67 kDa]) in transfected (T) K562 cells, respectively, while no visible expression was observed in Untransfected (UT) cells, 168h post transfection. β-actin is used as the loading positive control. (**D.i.)** Representative confocal images of K562-A*33 (T) and K562-A*24 (T) cells infected with *Toxoplasma* tachyzoites using a red/green invasion assay. External tachyzoites are stained green (SAG1), internal tachyzoites are stained red (GAP45) and host nuclei are DAPI stained-blue. (**D.ii.**) Quantification of % invasion (Total Red tachyzoites/Total Green tachyzoites*100) using the red/green assay. (**E.i.**) Representative 10X Giemsa stain images of A*33 and A*24 infected PBMCs at 24h, 48h, 72h, 120h and 168h P.I. (**E.ii.**) Bar graph showing percentage of live cells at various time points from E.i. Data is represented as Mean ± SD from 3 independent experiments. p ≤ 0.05 is marked as *, p ≤ 0.01 is marked as ** and p ≤ 0.001 is marked as ***.

**Supplementary Figure 2.**
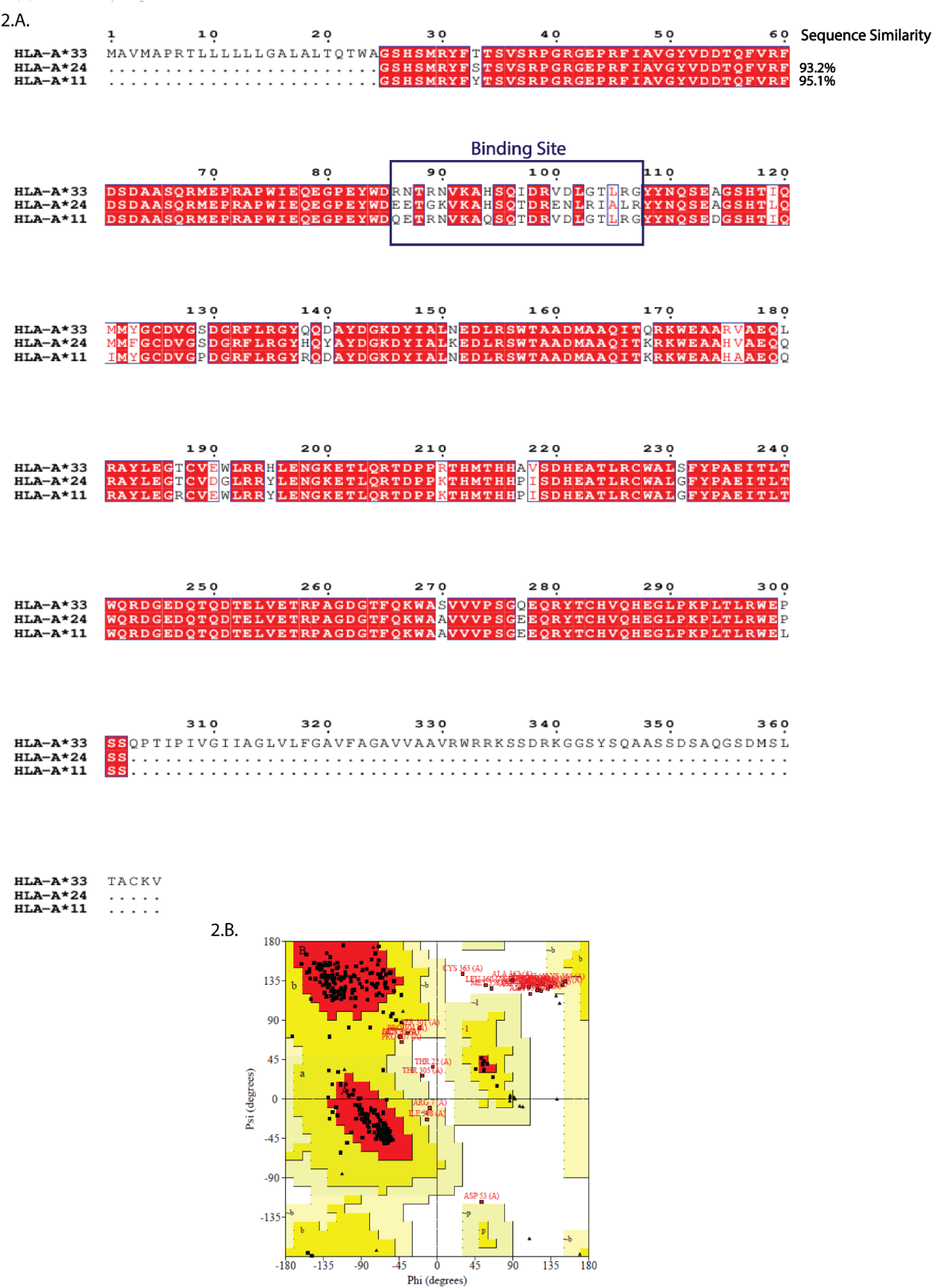
**(A.)** Multiple Sequence alignment of HLA-A*33 whole protein showing 95.1% sequence similarity with HLA-A*\**11 and 93.2% sequence similarity with HLA-A*24. The blue box highlights the binding site, i.e. residues 86 to 107 of A*33, with A*11 showing the maximum similarity in binding site residues (86.3%). *(**B.**)* Ramachandran plot of the selected A*33 model, illustrating the distribution of backbone dihedral angles. The plot shows 83.7% of residues are located in the core allowed (A, B, L) regions, while 7.2% are in the additional allowed (a, b, l, p) regions, indicating the structural stability of the model.

## Supplementary Tables

**Supplementary** Table 1: Excel Sheet of 92 alleles from NGS data of HbE/β-thalassemia patients

**Supplementary** Table 2: Excel Sheet of alleles from confirmatory Sanger sequencing of Healthy Controls

## References

[1] R. Colah, A. Gorakshakar, and A. Nadkarni, “Global burden, distribution and prevention of β-thalassemias and hemoglobin E disorders.,” Expert Rev. Hematol., vol. 3, no. 1, pp. 103–117, Feb. 2010, doi: 10.1586/ehm.09.74.

[2] D. Mohanty et al., “Prevalence of β-thalassemia and other haemoglobinopathies in six cities in India: a multicentre study.,” J. Community Genet., vol. 4, no. 1, pp. 33–42, Jan. 2013, doi: 10.1007/s12687-012-0114-0.

[3] A. Kattamis, G. L. Forni, Y. Aydinok, and V. Viprakasit, “Changing patterns in the epidemiology of β-thalassemia.,” Eur. J. Haematol., vol. 105, no. 6, pp. 692–703, Dec. 2020, doi: 10.1111/ejh.13512.

[4] S. S. Yadav, P. Panchal, and K. C. Menon, “Prevalence and Management of β-Thalassemia in India,” Hemoglobin, vol. 46, no. 1, pp. 27–32, 2022, doi: 10.1080/03630269.2021.2001346.

[5] P. Charoenkwan and A. Tantiworawit, “Treatment strategies for haemoglobin E thalassaemia.,” Lancet. Glob. Heal., vol. 10, no. 1, pp. e18–e19, Jan. 2022, doi: 10.1016/S2214-109X(21)00507-6.

[6] S. Uyoga et al., “Sickle cell anaemia and severe Plasmodium falciparum malaria: a secondary analysis of the Transfusion and Treatment of African Children Trial (TRACT).,” Lancet. Child Adolesc. Heal., vol. 6, no. 9, pp. 606–613, Sep. 2022, doi: 10.1016/S2352-4642(22)00153-5.

[7] K. Ayi, F. Turrini, A. Piga, and P. Arese, “Enhanced phagocytosis of ring-parasitized mutant erythrocytes: a common mechanism that may explain protection against falciparum malaria in sickle trait and beta-thalassemia trait.,” Blood, vol. 104, no. 10, pp. 3364–3371, Nov. 2004, doi: 10.1182/blood-2003-11-3820.

[8] K. Chotivanich et al., “Hemoglobin E: a balanced polymorphism protective against high parasitemias and thus severe P falciparum malaria.,” Blood, vol. 100, no. 4, pp. 1172–1176, Aug. 2002.

[9] A. V Hill et al., “Common west African HLA antigens are associated with protection from severe malaria.,” Nature, vol. 352, no. 6336, pp. 595–600, Aug. 1991, doi: 10.1038/352595a0.

[10] Z. Karakas et al., “THE FREQUENCY OF HLA-A, B AND DRB1 ALLELES IN PATIENTS WITH BETA THALASSEMIA,” Hematol. Transfus. Cell Ther., vol. 43, pp. S26–S27, 2021, doi: 10.1016/j.htct.2021.10.996.

[11] J. Yoon, “Acute Myeloid Leukemia Is a Disease Associated with HLA-C3,” Acta Haematol., vol. 133, no. 2, pp. 164–167, 2014, doi: 10.1159/000365436.

[12] T. Shiina, K. Hosomichi, H. Inoko, and J. K. Kulski, “The HLA genomic loci map: expression, interaction, diversity and disease.,” J. Hum. Genet., vol. 54, no. 1, pp. 15– 39, Jan. 2009, doi: 10.1038/jhg.2008.5.

[13] K. Hirayasu et al., “Evidence for natural selection on leukocyte immunoglobulin-like receptors for HLA class I in Northeast Asians.,” Am. J. Hum. Genet., vol. 82, no. 5, pp. 1075–1083, May 2008, doi: 10.1016/j.ajhg.2008.03.012.

[14] K. Ghosh, “Evolution and selection of human leukocyte antigen alleles by Plasmodium falciparum infection.,” Hum. Immunol., vol. 69, no. 12, pp. 856–860, Dec. 2008, doi: 10.1016/j.humimm.2008.08.294.

[15] A. V Hill et al., “Human leukocyte antigens and natural selection by malaria.,” *Philos. Trans. R. Soc. London. Ser. B*, Biol. Sci., vol. 346, no. 1317, pp. 379–385, Nov. 1994, doi: 10.1098/rstb.1994.0155.

[16] D. Meyer, V. R. C Aguiar, B. D. Bitarello, D. Y. C Brandt, and K. Nunes, “A genomic perspective on HLA evolution.,” Immunogenetics, vol. 70, no. 1, pp. 5–27, Jan. 2018, doi: 10.1007/s00251-017-1017-3.

[17] I. Lisovsky, G. Isitman, J. Bruneau, and N. F. Bernard, “Functional analysis of NK cell subsets activated by 721.221 and K562 HLA-null cells.,” J. Leukoc. Biol., vol. 97, no. 4, pp. 761–767, Apr. 2015, doi: 10.1189/jlb.4AB1014-499R.

[18] J. Sutherland, P. Mannoni, F. Rosa, D. Huyat, A. R. Turner, and M. Fellous, “Induction of the expression of HLA class I antigens on K562 by interferons and sodium butyrate.,” Hum. Immunol., vol. 12, no. 2, pp. 65–73, Feb. 1985, doi: 10.1016/0198-8859(85)90344-1.

[19] N. Hirano et al., “Efficient presentation of naturally processed HLA class I peptides by artificial antigen-presenting cells for the generation of effective antitumor responses.,” Clin. cancer Res. an Off. J. Am. Assoc. Cancer Res., vol. 12, no. 10, pp. 2967–2975, May 2006, doi: 10.1158/1078-0432.CCR-05-2791.

[20] C. M. Britten, R. G. Meyer, T. Kreer, I. Drexler, T. Wölfel, and W. Herr, “The use of HLA-A*0201-transfected K562 as standard antigen-presenting cells for CD8(+) T lymphocytes in IFN-gamma ELISPOT assays.,” J. Immunol. Methods, vol. 259, no. 1– 2, pp. 95–110, Jan. 2002, doi: 10.1016/s0022-1759(01)00499-9.

[21] I. J. Fuss, M. E. Kanof, P. D. Smith, and H. Zola, “Isolation of whole mononuclear cells from peripheral blood and cord blood.,” Curr. Protoc. Immunol., vol. Chapter 7, pp. 7.1.1–7.1.8, Apr. 2009, doi: 10.1002/0471142735.im0701s85.

[22] Q. Zhang, K. Hall, and M. Tally, “Partial purification of an IGF-II receptor/binding protein from the erythroleukemia cell line K562.,” Mol. Cell. Biochem., vol. 154, no. 1, pp. 47–54, Jan. 1996, doi: 10.1007/BF00248460.

[23] T. O. Artamonova, M. A. Khodorkovskiĭ, and A. S. Tsimokha, “[Mass spectrometric analysis of proteasomes affinity puirified from the human myelogenous leukemia cells K562].,” Bioorg. Khim., vol. 40, no. 6, pp. 720–734, 2014, doi: 10.1134/s1068162014060041.

[24] B. Mukherjee et al., “Antimony-resistant but not antimony-sensitive Leishmania donovani up-regulates host IL-10 to overexpress multidrug-resistant protein 1.,” Proc. Natl. Acad. Sci. U. S. A., vol. 110, no. 7, pp. E575–82, Feb. 2013, doi: 10.1073/pnas.1213839110.

[25] M. Wieczorek et al., “Major Histocompatibility Complex (MHC) Class I and MHC Class II Proteins: Conformational Plasticity in Antigen Presentation.,” Front. Immunol., vol. 8, p. 292, 2017, doi: 10.3389/fimmu.2017.00292.

[26] N. R. Shih et al., “HLA class I peptide polymorphisms contribute to class II DQβ0603:DQα0103 antibody specificity.,” Nat. Commun., vol. 15, no. 1, p. 609, Jan. 2024, doi: 10.1038/s41467-024-44912-0.

[27] O. Serçinoğlu and P. Ozbek, “Sequence-structure-function relationships in class I MHC: A local frustration perspective.,” PLoS One, vol. 15, no. 5, p. e0232849, 2020, doi: 10.1371/journal.pone.0232849.

[28] I. Hassan and F. Ahmad, “Structural diversity of class I MHC-like molecules and its implications in binding specificities.,” Adv. Protein Chem. Struct. Biol., vol. 83, pp. 223–270, 2011, doi: 10.1016/B978-0-12-381262-9.00006-9.

[29] X. C. Li and M. Raghavan, “Structure and function of major histocompatibility complex class I antigens.,” Curr. Opin. Organ Transplant., vol. 15, no. 4, pp. 499– 504, Aug. 2010, doi: 10.1097/MOT.0b013e32833bfb33.

[30] K. M. Musallam, M. D. Cappellini, V. Viprakasit, A. Kattamis, S. Rivella, and A. T. Taher, “Revisiting the non-transfusion-dependent (NTDT) vs. transfusion-dependent (TDT) thalassemia classification 10 years later.,” American journal of hematology, vol. 96, no. 2. United States, pp. E54–E56, Feb. 2021. doi: 10.1002/ajh.26056.

[31] M. B. Purner et al., “CD4-mediated and CD8-mediated cytotoxic and proliferative immune responses to Toxoplasma gondii in seropositive humans.,” Infect. Immun., vol. 64, no. 10, pp. 4330–4338, Oct. 1996, doi: 10.1128/iai.64.10.4330-4338.1996.

[32] K. Du et al., “Toxoplasma gondii infection induces cell apoptosis via multiple pathways revealed by transcriptome analysis.,” J. Zhejiang Univ. Sci. B, vol. 23, no. 4, pp. 315–327, Apr. 2022, doi: 10.1631/jzus.B2100877.

[33] D. Su et al., “Toxoplasma gondii infection regulates apoptosis of host cells via miR-185/ARAF axis.,” Parasit. Vectors, vol. 16, no. 1, p. 371, Oct. 2023, doi: 10.1186/s13071-023-05991-y.

[34] A. Hernández-de-Los-Ríos et al., “Influence of Two Major Toxoplasma Gondii Virulence Factors (ROP16 and ROP18) on the Immune Response of Peripheral Blood Mononuclear Cells to Human Toxoplasmosis Infection.,” Front. Cell. Infect. Microbiol., vol. 9, p. 413, 2019, doi: 10.3389/fcimb.2019.00413.

[35] C. S. Meira et al., “Cerebral and ocular toxoplasmosis related with IFN-γ, TNF-α, and IL-10 levels.,” Front. Microbiol., vol. 5, p. 492, 2014, doi: 10.3389/fmicb.2014.00492.

[36] R. T. Gazzinelli, F. T. Hakim, S. Hieny, G. M. Shearer, and A. Sher, “Synergistic role of CD4+ and CD8+ T lymphocytes in IFN-gamma production and protective immunity induced by an attenuated Toxoplasma gondii vaccine.,” J. Immunol., vol. 146, no. 1, pp. 286–292, Jan. 1991.

[37] Y. Suzuki, M. A. Orellana, R. D. Schreiber, and J. S. Remington, “Interferon-gamma: the major mediator of resistance against Toxoplasma gondii.,” Science, vol. 240, no. 4851, pp. 516–518, Apr. 1988, doi: 10.1126/science.3128869.

[38] C. R. Sturge and F. Yarovinsky, “Complex immune cell interplay in the gamma interferon response during Toxoplasma gondii infection.,” Infect. Immun., vol. 82, no. 8, pp. 3090–3097, Aug. 2014, doi: 10.1128/IAI.01722-14.

[39] L. D. Saffer, S. A. Long Krug, and J. D. Schwartzman, “The role of phospholipase in host cell penetration by Toxoplasma gondii,” Am. J. Trop. Med. Hyg., vol. 40, no. 2, p. 145—149, Feb. 1989, doi: 10.4269/ajtmh.1989.40.145.

[40] V. Sabaj, M. Galindo, D. Silva, L. Sandoval, and J. C. Rodríguez, “Analysis of Toxoplasma gondii surface antigen 2 gene (SAG2). Relevance of genotype I in clinical toxoplasmosis.,” Mol. Biol. Rep., vol. 37, no. 6, pp. 2927–2933, Jul. 2010, doi: 10.1007/s11033-009-9854-2.

[41] C. Lekutis, D. J. Ferguson, M. E. Grigg, M. Camps, and J. C. Boothroyd, “Surface antigens of Toxoplasma gondii: variations on a theme.,” Int. J. Parasitol., vol. 31, no. 12, pp. 1285–1292, Oct. 2001, doi: 10.1016/s0020-7519(01)00261-2.

[42] K. Winkler et al., “Changing the antigen binding specificity by single point mutations of an anti-p24 (HIV-1) antibody.,” J. Immunol., vol. 165, no. 8, pp. 4505–4514, Oct. 2000, doi: 10.4049/jimmunol.165.8.4505.

[43] J. E. Barnes, P. K. Lund-Andersen, J. S. Patel, and F. M. Ytreberg, “The effect of mutations on binding interactions between the SARS-CoV-2 receptor binding domain and neutralizing antibodies B38 and CB6.,” Sci. Rep., vol. 12, no. 1, p. 18819, Nov. 2022, doi: 10.1038/s41598-022-23482-5.

[44] R. Kronstein-Wiedemann et al., “K562 erythroleukemia line as a possible reticulocyte source to culture Plasmodium vivax and its surrogates.,” Exp. Hematol., vol. 82, pp. 8–23, Feb. 2020, doi: 10.1016/j.exphem.2020.01.012.

[45] T. G. Theander et al., “Cell-mediated immunity to Plasmodium falciparum infection: evidence against the involvement of cytotoxic lymphocytes.,” Scand. J. Immunol., vol. 28, no. 1, pp. 105–111, Jul. 1988, doi: 10.1111/j.1365-3083.1988.tb02421.x.

[46] L. M. Neri et al., “Erythropoietin-induced erythroid differentiation of K562 cells is accompanied by the nuclear translocation of phosphatidylinositol 3-kinase and intranuclear generation of phosphatidylinositol (3,4,5) trisphosphate.,” Cell. Signal., vol. 14, no. 1, pp. 21–29, Jan. 2002, doi: 10.1016/s0898-6568(01)00224-8.

[47] N. Uchida et al., “High-level embryonic globin production with efficient erythroid differentiation from a K562 erythroleukemia cell line.,” Exp. Hematol., vol. 62, pp. 7–16.e1, Jun. 2018, doi: 10.1016/j.exphem.2018.02.007.

[48] K. E. Lyke et al., “Association of HLA alleles with Plasmodium falciparum severity in Malian children.,” Tissue Antigens, vol. 77, no. 6, pp. 562–571, Jun. 2011, doi: 10.1111/j.1399-0039.2011.01661.x.

[49] A. O’Donnell et al., “Interaction of malaria with a common form of severe thalassemia in an Asian population.,” Proc. Natl. Acad. Sci. U. S. A., vol. 106, no. 44, pp. 18716– 18721, Nov. 2009, doi: 10.1073/pnas.0910142106.

[50] J. Kuesap, W. Chaijaroenkul, K. Rungsihirunrat, K. Pongjantharasatien, and K. Na-Bangchang, “Coexistence of Malaria and Thalassemia in Malaria Endemic Areas of Thailand.,” Korean J. Parasitol., vol. 53, no. 3, pp. 265–270, Jun. 2015, doi: 10.3347/kjp.2015.53.3.265.

[51] M. Ravenhall et al., “Novel genetic polymorphisms associated with severe malaria and under selective pressure in North-eastern Tanzania.,” PLoS Genet., vol. 14, no. 1, p. e1007172, Jan. 2018, doi: 10.1371/journal.pgen.1007172.

[52] J. L. Carson, “Blood transfusion and risk of infection: new convincing evidence.,” JAMA, vol. 311, no. 13. United States, pp. 1293–1294, Apr. 2014. doi: 10.1001/jama.2014.2727.

[53] L. T. Goodnough, “Risks of blood transfusion.,” Crit. Care Med., vol. 31, no. 12 Suppl, pp. S678-86, Dec. 2003, doi: 10.1097/01.CCM.0000100124.50579.D9.

[54] M. P. Busch, S. H. Kleinman, and G. J. Nemo, “Current and emerging infectious risks of blood transfusions.,” JAMA, vol. 289, no. 8, pp. 959–962, Feb. 2003, doi: 10.1001/jama.289.8.959.

[55] E. P. Dellinger and D. A. Anaya, “Infectious and immunologic consequences of blood transfusion.,” Crit. Care, vol. 8 Suppl 2, no. Suppl 2, pp. S18-23, 2004, doi: 10.1186/cc2847.

[56] S. Medhasi and N. Chantratita, “Human Leukocyte Antigen (HLA) System: Genetics and Association with Bacterial and Viral Infections.,” J. Immunol. Res., vol. 2022, p. 9710376, 2022, doi: 10.1155/2022/9710376.

[57] F. Prugnolle, A. Manica, M. Charpentier, J. F. Guégan, V. Guernier, and F. Balloux, “Pathogen-driven selection and worldwide HLA class I diversity.,” Curr. Biol., vol. 15, no. 11, pp. 1022–1027, Jun. 2005, doi: 10.1016/j.cub.2005.04.050.

[58] S. C. Gilbert et al., “Association of malaria parasite population structure, HLA, and immunological antagonism.,” Science, vol. 279, no. 5354, pp. 1173–1177, Feb. 1998, doi: 10.1126/science.279.5354.1173.

[59] W. N. A. M. Asuar et al., “β-Thalassemia Major Association with Human Leukocyte Antigen (HLA) Genes in the Malaysian Population,” Hemoglobin, vol. 43, no. 6, p. 345, 2019, doi: 10.1080/03630269.2020.1717134.

[60] A. T. Nguyen, C. Szeto, and S. Gras, “The pockets guide to HLA class I molecules.,” Biochem. Soc. Trans., vol. 49, no. 5, pp. 2319–2331, Nov. 2021, doi: 10.1042/BST20210410.

[61] C. McMurtrey et al., “Toxoplasma gondii peptide ligands open the gate of the HLA class I binding groove.,” Elife, vol. 5, Jan. 2016, doi: 10.7554/eLife.12556.

[62] K. El Bissati et al., “Adjuvanted multi-epitope vaccines protect HLA-A*11:01 transgenic mice against Toxoplasma gondii.,” JCI insight, vol. 1, no. 15, p. e85955, Sep. 2016, doi: 10.1172/jci.insight.85955.

[63] N. I. Cardona, D. M. Moncada, and J. E. Gómez-Marin, “A rational approach to select immunogenic peptides that induce IFN-γ response against Toxoplasma gondii in human leukocytes.,” Immunobiology, vol. 220, no. 12, pp. 1337–1342, Dec. 2015, doi: 10.1016/j.imbio.2015.07.009.

[64] A. Jain et al., “IndiGenomes: a comprehensive resource of genetic variants from over 1000 Indian genomes,” Nucleic Acids Res., vol. 49, no. D1, pp. D1225–D1232, 2020, doi: 10.1093/nar/gkaa923.

[65] S. Singh, A. Munawwar, S. Rao, S. Mehta, and N. K. Hazarika, “Serologic prevalence of Toxoplasma gondii in Indian women of child bearing age and effects of social and environmental factors.,” PLoS Negl. Trop. Dis., vol. 8, no. 3, p. e2737, Mar. 2014, doi: 10.1371/journal.pntd.0002737.

## References

1. Wieczorek, M., et al. Major Histocompatibility Complex (MHC) Class I and MHC Class II Proteins: Conformational Plasticity in Antigen Presentation. Front. Immunol. 8, 292 (2017).

2. Shih, N. R., et al. HLA class I peptide polymorphisms contribute to class II DQβ 0603:DQα0103 antibody specificity. Nat. Commun. 15, 609 (2024).

3. Li, X. C. & Raghavan, M. Structure and function of major histocompatibility complex class I antigens. Curr. Opin. Organ Transplant. 15, 499–504 (2010).

4. Serçinoğlu, O. & Ozbek, P. Sequence-structure-function relationships in class I MHC: A local frustration perspective. PLoS One 15, e0232849 (2020).

5. Hassan, I. & Ahmad, F. Structural diversity of class I MHC-like molecules and its implications in binding specificities. Adv. Protein Chem. Struct. Biol. 83, 223–270 (2011).

6. Barker, D. J. et al. The IPD-IMGT/HLA Database. Nucleic Acids Res. 51, D1053– D1060 (2023).

7. Madeira, F., et al. The EMBL-EBI Job Dispatcher sequence analysis tools framework in 2024. Nucleic Acids Res. 52, W521–W525 (2024).

8. Jumper, J., et al. Highly accurate protein structure prediction with AlphaFold. Nature 596, 583–589 (2021).

9. Mirdita, M., et al. ColabFold: making protein folding accessible to all. Nat. Methods 19, 679–682 (2022).

10. Humphrey, W., Dalke, A. & Schulten, K. VMD: visual molecular dynamics. J. Mol. Graph. 14, 27–28,33-38 (1996).

11. Gajria, B., et al. ToxoDB: an integrated Toxoplasma gondii database resource. Nucleic Acids Res. 36, D553–6 (2008).

12. Dominguez, C., Boelens, R. & Bonvin, A. M. J. J. HADDOCK: a protein-protein docking approach based on biochemical or biophysical information. J. Am. Chem. Soc. 125, 1731–1737 (2003).

13. van Zundert, G. C. P., et al. The HADDOCK2.2 Web Server: User-Friendly Integrative Modeling of Biomolecular Complexes. J. Mol. Biol. 428, 720–725 (2016).

